# Single-nucleus RNA sequencing identifies transcriptomic signatures of alcohol use disorder in the human ventral tegmental area

**DOI:** 10.64898/2026.05.15.26353305

**Authors:** Sofiya Patra, Ji Sun Koo, Abhyuday Singh Parihar, Chao Zhang, Huiping Zhang

**Author notes:** Correspondence: Huiping Zhang, Ph.D., Department of Psychiatry and Department of Medicine (Biomedical Genetics), Boston University Chobanian and Avedisian School of Medicine, Boston MA02118, USA, Chao Zhang, Ph.D., Department of Medicine (Computational Biomedicine), Boston University Chobanian and Avedisian School of Medicine, Boston MA02118, USA.

## Abstract

**Background:** Alcohol use disorder (AUD) is associated with altered gene expression across diverse cell types in reward-related brain regions, including the ventral tegmental area (VTA), which is rich in dopaminergic neurons. The VTA plays a central role in reward processing, learning, and memory; however, cell type-specific gene expression changes within the VTA remain uncharacterized.

**Methods:** We applied single-nucleus RNA sequencing (snRNA-seq) to profile transcriptomic alterations associated with AUD in the VTA. Postmortem VTA samples from four individuals of European ancestry [two with AUD (one male, one female) and two matched controls (one male, one female)] were analyzed using the 10X Genomics Chromium Fixed RNA Profiling protocol. Differentially expressed genes (DEGs) were identified using Seurat, and enriched KEGG pathways was assessed by gene set enrichment analysis.

**Results:** Nuclei were classified into six major cell types: astrocytes, endothelial cells, mature neurons, microglia, oligodendrocytes, and oligodendrocyte precursor cells (OPCs). At thresholds of *P* < 0.05 and |fold change| > 2.0, we identified 547 DEGs in astrocytes, 727 DEGs in endothelial cells, 715 DEGs in mature neurons, 421 DEGs in microglia, 263 DEGs in oligodendrocytes, and 432 DEGs in OPCs. DEGs across VTA cell types were enriched for pathways related to mitochondrial function, neurodegeneration, and synaptic signaling. Notably, DEGs in mature neurons were enriched for addiction-related pathways. Further subdivision of mature neurons into dopaminergic, GABAergic, glutamatergic, and unclassified subtypes revealed 526, 930, 896, and 569 DEGs, respectively. Neuronal DEGs indicate a convergence on mitochondrial/oxidative phosphorylation and neurodegeneration-related pathways across subtypes, whereas addiction- and synapse-related pathways show dopaminergic neuron-specific enrichment.

**Conclusions:** This study provides the first cell type-resolved transcriptomic profiling of the human VTA, revealing AUD-associated gene expression alterations across neuronal, glial, and endothelial cells. The observed cell type-specific changes in synaptic plasticity and addiction-related genes offer new insights into molecular mechanisms underlying AUD pathophysiology.

## INTRODUCTION

Alcohol use disorder (AUD) is a chronic, relapsing neuropsychiatric condition characterized by compulsive alcohol consumption and adverse health consequences (Koob and Volkow, 2016). Understanding gene expression alterations associated with AUD within key addiction-related brain regions is essential for developing targeted therapeutic strategies to treat the disorder. Prior transcriptomic studies using microarray and bulk RNA sequencing (RNA-seq) have revealed widespread gene expression changes across multiple brain areas implicated in addiction and alcohol response, including the amygdala (AMY), caudate nucleus (CN), cerebellum (CRB), hippocampus (HIPPO), nucleus accumbens (NAc), prefrontal cortex (PFC), putamen (PUT), and ventral tegmental area (VTA) (Besong et al., 2024, Farris et al., 2015, Flatscher-Bader et al., 2005, Lewohl et al., 2000, Lim et al., 2021, Liu et al., 2004, Liu et al., 2006, Mayfield et al., 2002, McClintick et al., 2013, Zhang et al., 2014). These studies have highlighted dysregulation in pathways involving synaptic plasticity, neuroinflammation, and metabolic processes (Mayfield et al., 2013, Ponomarev et al., 2012). However, bulk RNA analyses average gene expression signals across heterogeneous cell populations, potentially masking critical cell type-specific transcriptional changes relevant to AUD neuropathology.

Single-cell RNA sequencing (scRNA-seq) has emerged as a powerful technology enabling transcriptomic profiling at cellular resolution, providing detailed insights into cell type diversity and transcriptional states (Yao et al., 2021). Nonetheless, its application to frozen postmortem tissue samples is limited because intact whole cells are often difficult to isolate due to membrane disruption and RNA degradation after death, especially in the cytoplasm where most mRNA resides. By contrast, single-nucleus RNA sequencing (snRNA-seq) profiles RNA within isolated nuclei, predominantly capturing nuclear RNA and nascent transcripts. This method is well-suited to frozen or archived postmortem tissue samples, mitigating the impact of cytoplasmic RNA degradation and enabling reliable transcriptomic data recovery when fresh tissue is rarely available (Grindberg et al., 2013, Lake et al., 2016).

Recent snRNA-seq studies have revealed cell type-specific transcriptomic alterations in addiction-relevant brain regions, such as the prefrontal cortex (Brenner et al., 2020) and nucleus accumbens (van den Oord et al., 2023), from postmortem AUD brains. These investigations demonstrated distinct gene expression changes among neuronal and glial subpopulations linked to alcohol dependence, advancing our understanding of the cellular heterogeneity underpinning addictive behaviors. However, snRNA-seq analysis of the VTA, one of the brain’s core reward regions, has not yet been reported for AUD postmortem samples.

The VTA contains dopaminergic neurons critical for motivation, reinforcement learning, and addiction-related behaviors (Brenner et al., 2020). Chronic alcohol exposure induces molecular and functional adaptations in the VTA that contribute to AUD persistence (Koob, 2008). Our previous bulk RNA-seq study of human postmortem VTA tissue demonstrated that AUD-associated microRNA-mRNA networks regulate key biological pathways, including *Opioid Signaling*, *Neuronal CREB Signaling*, and *IL-8 signaling* (Lim et al., 2021). Thus, investigating the VTA at single-nucleus resolution provides a unique opportunity to uncover cell type-specific transcriptomic changes underlying AUD pathophysiology in this critical reward center.

Here, we present a pilot snRNA-seq analysis of the human VTA from individuals with AUD, delineating cell type-specific gene expression alterations. Our findings advance mechanistic understanding of AUD within the brain’s reward circuitry and identify potential molecular targets for therapeutic intervention.

## MATERIALS AND METHODS

### Human postmortem brain tissue samples

Freshly frozen ventral tegmental area (VTA) tissue samples were obtained from the New South Wales Brain Tissue Resource Centre (NSWBTRC), Australia. These samples were previously used in our transcriptomic study on alcohol use disorder (AUD) (project approval #: PID409) (Besong et al., 2024, Lim et al., 2021) and had been stored at -80°C in our lab. For the current single-nucleus RNA sequencing (snRNA-seq) study, we selected four VTA tissue samples: one male AUD patient, one female AUD patient, one male control, and one female control. All subjects were European Australians without a history of illicit drug abuse or major psychotic disorders (schizophrenia, bipolar disorder) as defined by DSM-IV criteria (American Psychiatric Association, 1994). Control subjects had no history of AUD. Cases and controls were matched by age, postmortem interval (PMI), tissue pH, brain weight, and cause of death (Table S1).

### Nuclei isolation from frozen postmortem brain tissues

Nuclei were isolated from postmortem VTA tissue samples following the Tissue Fixation & Dissociation protocol for Chromium Fixed RNA Profiling (CG000553 Rev B) (10X Genomics, Pleasanton, CA, USA) with minor modifications. Tissue fixation was performed using the Chromium Next GEM RNA Profiling Sample Fixation Kit (PN-1000414) (10X Genomics, Pleasanton, CA, USA). All procedures were conducted on dry ice or ice, with centrifugation at 500 × g for 5 min at 4°C (swinging-bucket rotor). Approximately 20-50 mg of frozen VTA tissue was placed on a pre-chilled glass petri dish on dry ice and finely minced with pre-chilled blades and spatulas. The minced tissue powder was suspended in Fixation Buffer at a ratio of 1 ml per 25 mg of tissue using a wide-bore pipette, followed by incubation at 4°C for 16-24 hours. After fixation, nuclei were pelleted by centrifugation, washed with 2 ml cold PBS, and resuspended in 1 ml pre-chilled Tissue Resuspension Buffer.

For dissociation, after removing the supernatant (Tissue Resuspension Buffer), 2 ml of pre-warmed (37°C) Dissociation Solution was added, and the tissue was transferred to a gentleMACS™ C Tube (Miltenyi Biotec, Auburn, CA, USA). Dissociation was performed on a gentleMACS Octo Dissociator with Heaters (Miltenyi Biotec) using the gentleMACS Program: (1) temp ON; (2) spin 50 rpm for 20 min; (3) spin 1000 rpm for 30 sec; (4) spin −1000 rpm for 30 sec; (5) end. Progress of nucleus dissociation was monitored by counting a 10 μl aliquot. The resulting nucleus suspension was filtered through a 30 μm filter to remove debris and undissociated tissue. Nuclei were pelleted by centrifugation, resuspended in 1 ml pre-chilled Quenching Buffer, and nuclear density and viability were assessed by AO/PI staining. Quality control metrics for nuclei extracted from the four VTA tissue samples are provided in Table S2.

### Chromium Fixed RNA Profiling library construction and sequencing

Gene expression libraries were constructed using the Chromium Fixed RNA Profiling Reagent Kits for Multiplexed Samples (10X Genomics, Pleasanton, CA, USA). Each fixed nucleus sample was hybridized with a unique probe set containing a sample-specific barcode. Four nucleus samples, each labeled with distinct probe barcodes (BC001-BC004), were pooled and loaded onto a Chromium Next GEM Chip Q for Gel Beads-in-Emulsion (GEM) generation. Following GEM formation, ligation and sample barcoding were performed *via* thermal cycling. Subsequently, GEMs were broken, and the ligated products were purified and pre-amplified by indexing PCR. The final libraries contained P5 and P7 sequencing priming sites.

Library quantification was performed using KAPA Library Quantification Kits (Roche, Indianapolis, IN, USA), and size distribution was assessed with Agilent Bioanalyzer High Sensitivity chips (Agilent Technologies, Santa Clara, CA, USA). The libraries exhibited a typical 10X Flex profile, with an average size of approximately 260 bp and 84.4% of fragments ranging between 150 and 450 bp (Figure S1), meeting sequencing quality standards. Sequencing was carried out on an Illumina NextSeq 2000 at the Boston University Medical Campus Microarray & Sequencing Core. Libraries were run across two lanes with paired-end reads. The run generated over 1.4 billion reads, with 95.16% of bases above Q30 quality and a 0.26% error rate. Quality control metrics for the snRNA-seq data are summarized in Table S2. The experimental workflow is illustrated in Figure S2.

### snRNA-seq data processing and analysis

Raw FASTQ files for four pooled samples were demultiplexed and aligned to the human reference genome (GRCh38/hg38) using CellRanger v8.0.1 (Zheng et al., 2017). Downstream analyses were conducted in R (v4.4.0) using the Seurat package (v5.3.0) (Hao et al., 2024). For each sample, filtered Seurat objects were generated by retaining nuclei with at least 200 detected genes and genes expressed in at least three nuclei. Quality control metrics, including detected features (or genes), counts, percentage of mitochondrial RNA (mtRNA), and percentage of ribosomal RNA (rRNA), were visualized (Figure S3a). Filtering criteria included nuclei with > 200 features, < 30,000 counts, mtRNA < 10%, and rRNA < 5%. The quality control metrics after filtering are shown in Figure S3b. Filtered nuclei were normalized using NormalizeData() to adjust for sequencing depth and technical variability. Highly variable genes (HVGs) were identified using FindVariableFeatures() to select the top 2,000 genes. The data were scaled *via* ScaleData(), followed by dimensionality reduction using RunPCA(). Subsequent steps included FindNeighbors(), FindClusters(), and RunUMAP() using top 10 principal components (PCs). Potential doublets were predicted with DoubletFinder (McGinnis et al., 2019). As negligible doublets were detected (Figure S4), no nuclei were removed. Results from unintegrated clustering analyses of all samples, as well as analyses stratified by sample identity and subject gender, are presented in Figure S5a, whereas Harmony clustering results are shown in Figure S5b.

Seurat objects from individual samples were merged and jointly processed through normalization, HVGs identification, scaling, principal component analysis (PCA), uniform manifold approximation and projection (UMAP), and clustering. Data integration was performed using Harmony through Seurat’s IntegrateLayers() function (Korsunsky et al., 2019). The final UMAP embedding was computed on 20 principal components (PCs), and clustering at a resolution of 0.05 identified six broad cell-type clusters (Figure 1a). These clusters were annotated using FindAllMarkers() combined with automated annotation *via* scType (Ianevski et al., 2022, Nader et al., 2024).

**Figure 1.**
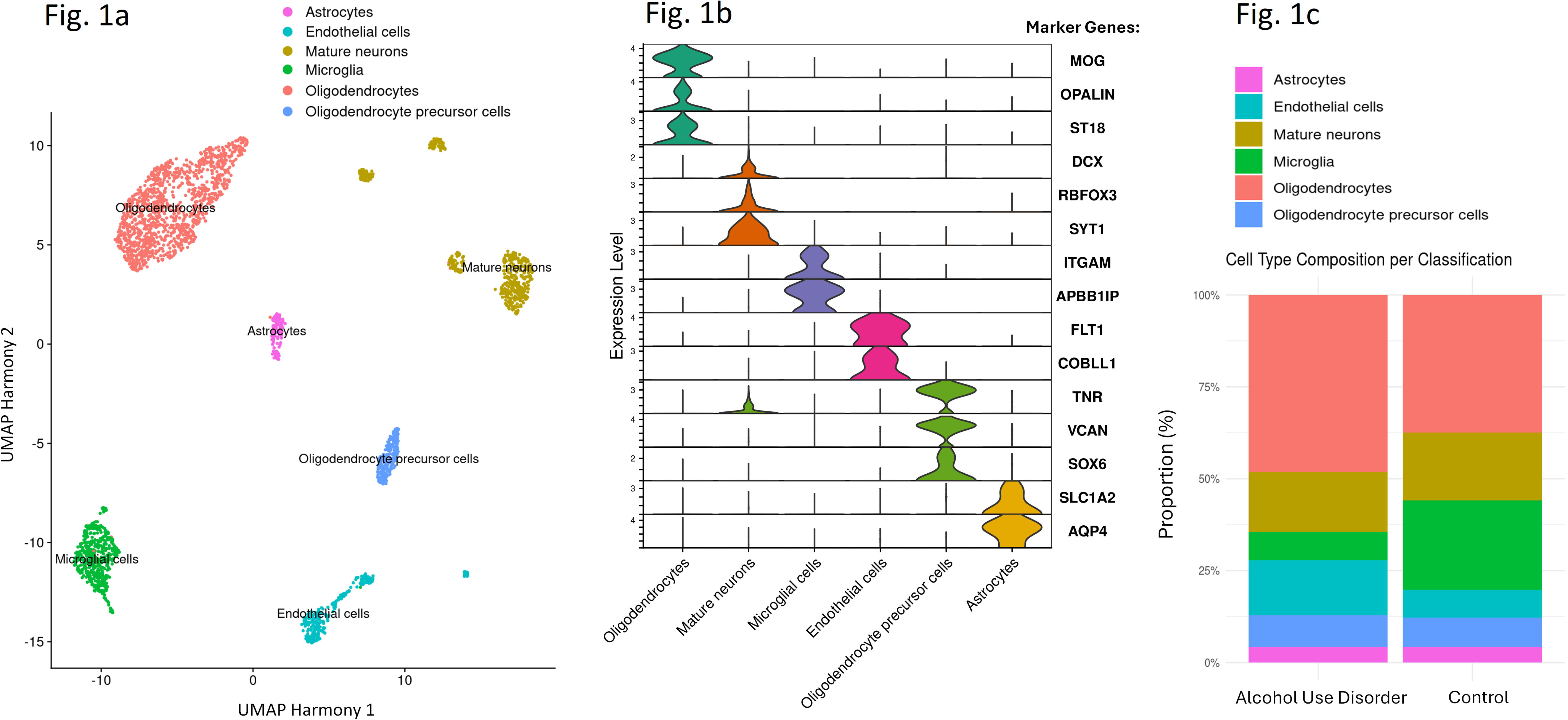
(a) Brain cell type identification from snRNA-seq data. (b) Violin plots showing cell type-specific marker gene expression. (c) Cell type proportions stratified by AUD status.

### Brain cell type-specific analysis

Six major cell-type subsets, including astrocytes, endothelial cells, mature neurons, microglia, oligodendrocytes, and oligodendrocyte precursor cells (OPCs) (Figure 1a), were identified based on previously published markers as described above. Differential gene expression between case and control groups was assessed with FindMarkers(). Marker expression and distribution were visualized using FeaturePlot() and VlnPlot(). Volcano plots of differentially expressed genes (DEGs) were generated with EnhancedVolcano (Blighe et al., 2025). Shared DEG counts across cell types were visualized using an UpSet plot (Lex et al., 2014). Pathway enrichment analysis was conducted with the ClusterProfiler package (v3.21), utilizing the enrichKEGG() function (Yu et al., 2012). DEGs with a *P*-value < 0.05 and absolute log2-based fold change (|log_2_FC|) > 1.0 were included for KEGG pathway enrichment analysis, applying a significance cutoff of *P*_adj_ < 0.05.

For neuron subtype analysis, mature neurons were further classified into dopaminergic, GABAergic, glutamatergic, and unclassified subtypes. Differential expression and functional annotation of DEGs between case and control groups were performed following the same workflow described above.

## RESULTS

### Brain cell type identification and composition differences by AUD status

Clustering analysis using Seurat identified six distinct brain cell types (astrocytes, endothelial cells, mature neurons, microglia, oligodendrocytes, and OPCs) (Figure 1a) based on the expression of specific marker genes (Figure 1b). Cell type compositions in postmortem VTA tissue from AUD and control subjects are shown in Figure 1c. AUD cases showed a higher proportion of endothelial cells and oligodendrocytes compared to controls [endothelial cells: 15.1% (case) vs. 7.5% (control); oligodendrocytes: 47.8% (case) vs. 37.7% (control)], whereas the proportion of microglia was lower in AUD cases (7.5%) than in controls (25.0%). The proportions of astrocytes, mature neurons, and OPCs were similar between the case and control groups. Among the six brain cell types, oligodendrocytes were the most abundant in the VTA of both AUD and control subjects (Figure 1c).

### Differentially expressed genes in six VTA cell types of AUD subjects

Differential expression analysis was performed separately for each cell type to compare AUD and control groups. Results are displayed as volcano plots in Figure 2. Using thresholds of unadjusted *P* < 0.05 and absolute log2-based fold change (|log_2_FC|) > 2.0, we identified DEGs as follows: 547 in astrocytes [103 (18.8%) upregulated, 444 (81.2%) downregulated]; 727 in endothelial cells [429 (59.0%) upregulated, 298 (41.0%) downregulated]; 715 in mature neurons [169 (23.6%) upregulated, 546 (76.4%) downregulated]; 421 in microglia [175 (41.6%) upregulated, 246 (58.4%) downregulated]; 263 in oligodendrocytes [54 (20.5%) upregulated, 209 (79.5%) downregulated]; and 432 in OPCs [255 (59.0%) upregulated, 177 (41.0%) downregulated]. Overall, alcohol exposure resulted in a higher number of downregulated genes in astrocytes, mature neurons, microglia, and oligodendrocytes, but more upregulated genes in endothelial cells and OPCs.

**Figure 2.**
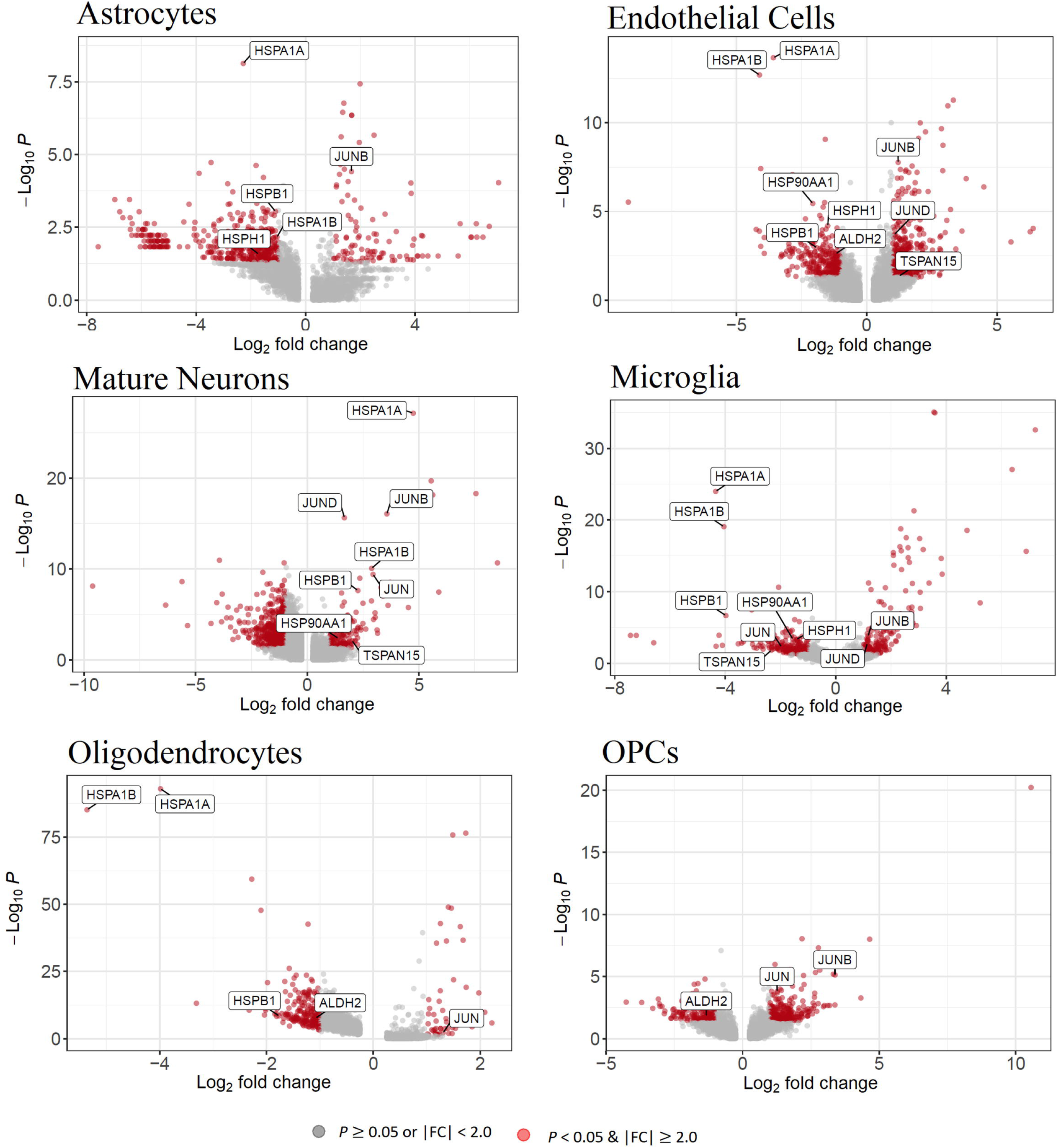
Volcano plots of differential gene expression across six VTA tissue cell types.

An UpSet plot (Figure 3a) illustrates the number of shared and unique DEGs (*P* < 0.05 & |log_2_FC| > 2.0) among the six cell types. The majority of DEGs identified in each cell type were specific to that cell type (unique DEGs in astrocytes: 399/547 = 72.9%; unique DEGs in endothelial cells: 573/727 = 78.8%; unique DEGs in mature neurons: 598/715 = 83.6%; unique DEGs in microglia: 307/421 = 72.9%; unique DEGs in oligodendrocytes: 169/263 = 64.2%; unique DEGs in OPCs: 322/432 = 74.5%). DEGs shared by more than three cell types are listed in Table 1. Notably, the acetaldehyde dehydrogenase 2 gene (*ALDH2*) was downregulated in three cell types: endothelial cells (*P* = 0.002, log_2_FC = -1.12), oligodendrocytes (*P* = 2.1×10^-8^, log_2_FC = -1.08), and OPCs (*P* = 0.015, log_2_FC = -1.33). The JUN gene, which encodes the c-Jun protein and functions as an immediate-early gene (IEG) critical for the brain’s response to addictive drugs (Zhou et al., 2014), was differentially expressed in four cell types: microglial cells (*P* = 0.005, log_2_FC = -1.96), mature neurons (*P* = 3.8×10^-10^, log_2_FC = 2.94), oligodendrocytes (*P* = 0.007, log_2_FC = 1.30), and OPCs (*P* = 0.0002, log_2_FC = 1.26). Additionally, three heat shock protein genes (*HSPA1A*, *HSPA1B*, and *HSPB1*) showed differential expression across five cell types: astrocytes, endothelial cells, microglial cells, mature neurons, and oligodendrocytes (Figure 2).

**Figure 3.**
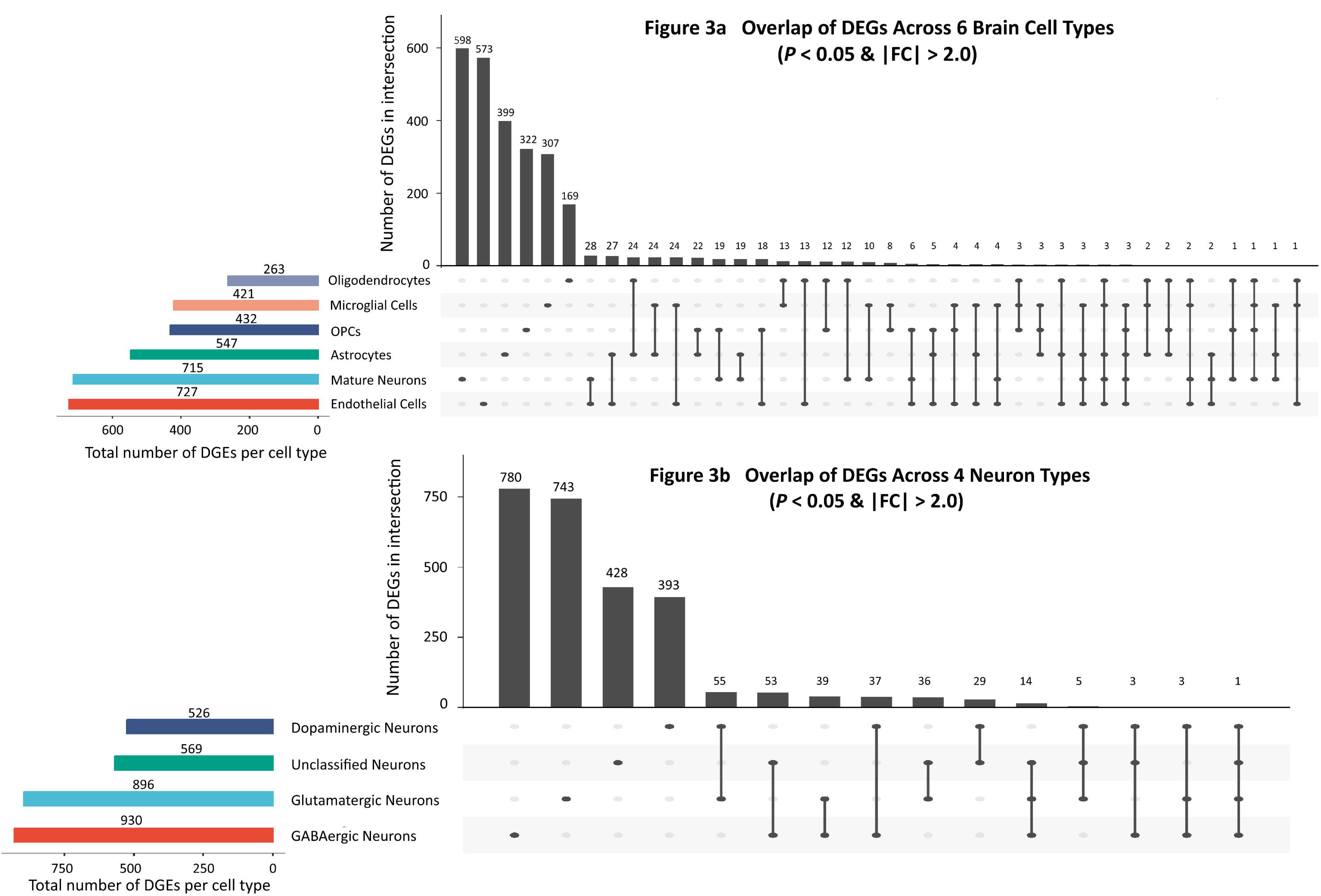
Upset plots depicting the numbers of shared DEGs among six VTA tissue cell types (a) and among four VTA neuronal subtypes (b).

**Table 1.**
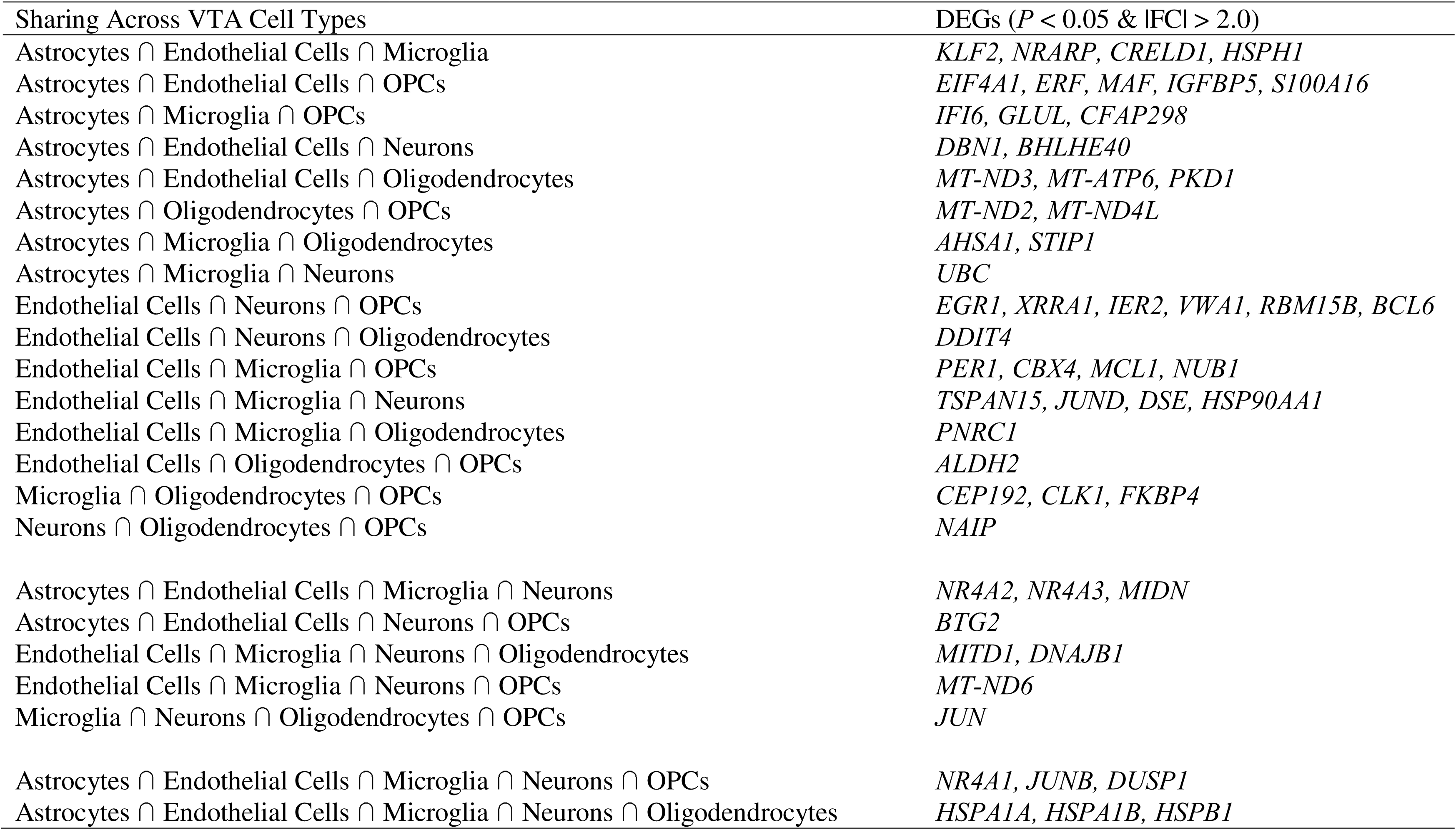
Differentially expressed genes (DEGs) shared across more than three brain cell types in the ventral tegmental area (VTA) of alcohol use disorder (AUD) subjects.

### Neuronal cell type identification and composition differences by AUD status

Neuronal cells were further subdivided into subtypes using clustering analysis (Figure 4a). Four neuronal subtypes were identified based on marker gene expression: glutamatergic (*SLC17A6*, *SLC17A7*, *CAMK2A*, *GRIK2*), dopaminergic (*TH*, *SLC6A3*, *SLC18A2*), GABAergic (*GAD1*, *GAD2*, *SLC32A1*, *GABRA1*), and unclassified neurons (Figure 4b). The composition of these neuronal subtypes in VTA tissue from AUD and control subjects is shown in Figure 4c. Compared to controls, AUD cases exhibited a lower proportion of glutamatergic neurons [36.4% (AUD) vs. 51.3% (Control)] and a higher proportion of unclassified neurons [36.9% (AUD) vs. 20.4% (Control)]. The proportions of dopaminergic and GABAergic neurons were comparable between the two groups.

**Figure 4.**
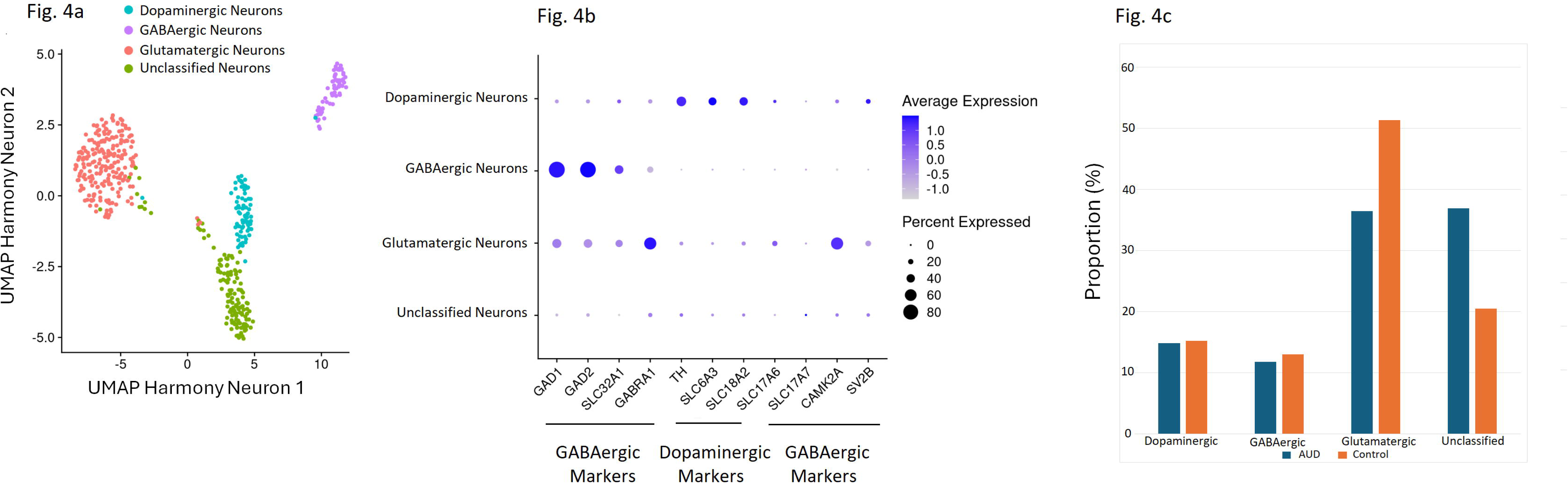
(a) UMAP showing four neuronal subtypes. (b) Marker gene expression. (c) Subtype proportions by AUD status.

### Differentially expressed genes in four neuronal subtypes of VTA Tissue of AUD subjects

Differential expression in the four neuronal subtypes is shown as volcano plots in Figure 5. Using thresholds of unadjusted *P* < 0.05 and |log_2_FC| > 2.0, we identified the following DEGs: 526 in dopaminergic neurons [324 (61.6%) upregulated, 202 (38.4%) downregulated]; 930 in GABAergic neurons [60 (6.5%) upregulated, 870 (93.5%) downregulated]; 896 in glutamatergic neurons [552 (61.6%) upregulated, 344 (38.4%) downregulated]; and 569 in unclassified neurons [247 (43.4%) upregulated, 322 (56.6%) downregulated]. Overall, alcohol exposure led to more upregulated genes in dopaminergic and glutamatergic neurons, whereas GABAergic and unclassified neurons showed more downregulated genes.

**Figure 5.**
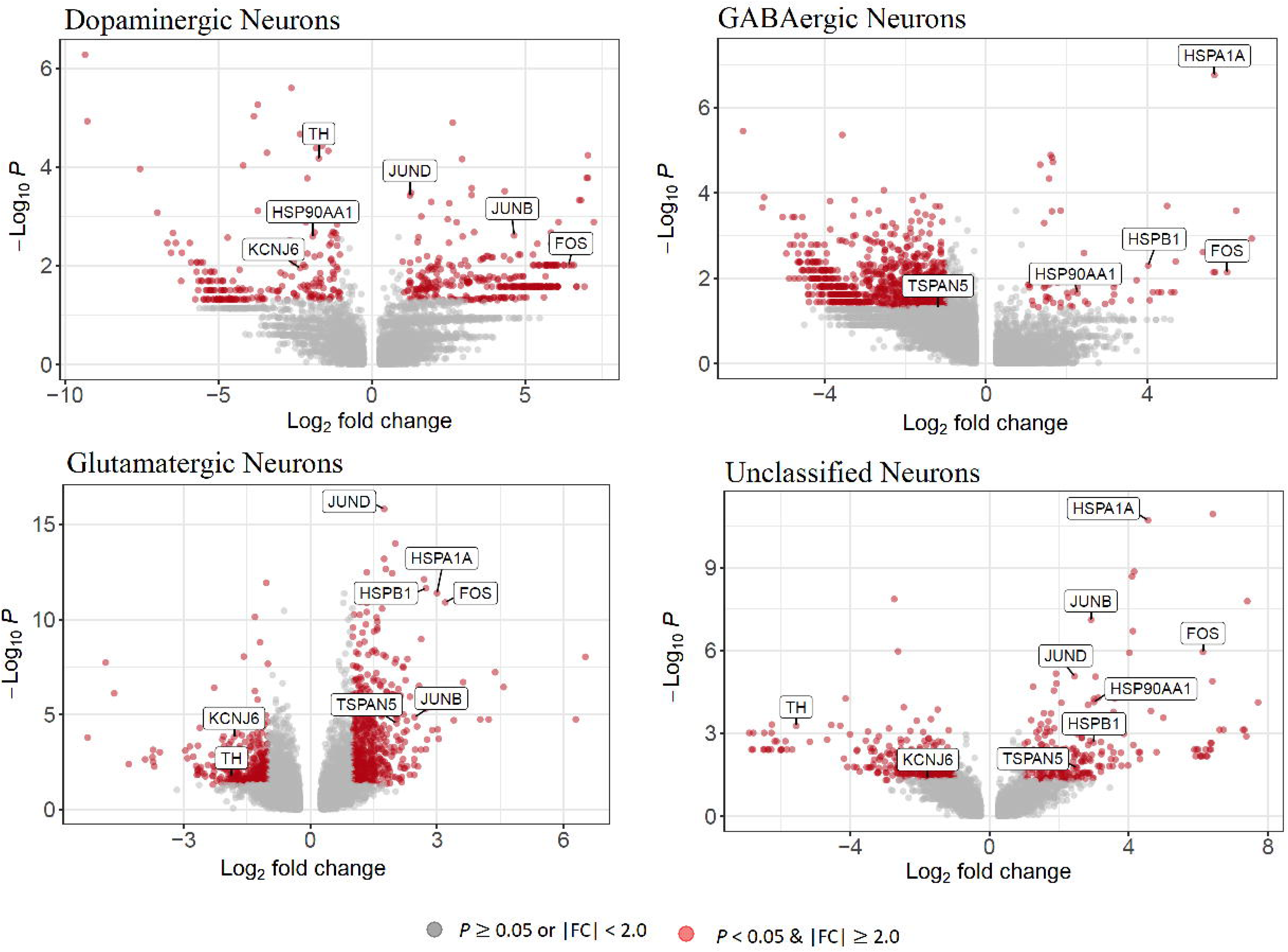
Volcano plots of differential gene expression across four VTA neuronal subtypes.

An Upset plot (Figure 3b) illustrates the number of shared and unique DEGs (*P* < 0.05 & |log_2_FC| > 2.0) among the four neuronal subtypes. Most DEGs were unique to each subtype, with the following proportions of unique DEGs: dopaminergic neurons, 393/526 (74.7%); GABAergic neurons, 780/930 (83.9%); glutamatergic neurons, 743/896 (82.9%); and unclassified neurons, 428/569 (75.2%). DEGs shared by more than three neuronal subtypes are presented in Table 2. Notably, the FOS gene, which produces a truncated splice variant, ΔFosB, known to act as a “molecular switch” mediating long-term neural and behavioral changes linked to addiction (McReynolds et al., 2018), was differentially expressed across all four neuronal subtypes.

**Table 2.**
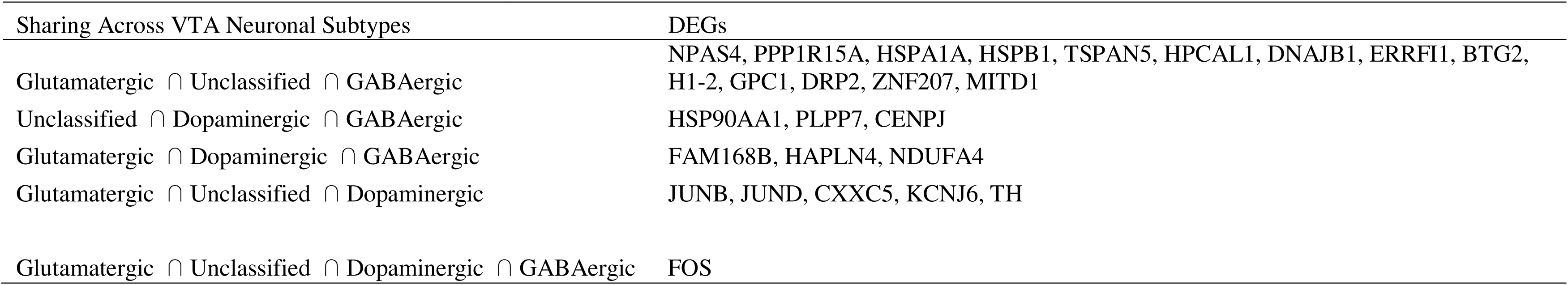
Differentially expressed genes (DEGs) shared across more than three neuronal subtypes in the ventral tegmental area (VTA) of alcohol use disorder (AUD) Subjects.

### Functional enrichment of DEGs in six VTA cell types

We performed KEGG pathway analysis on DEGs from six VTA cell types (DEGs defined as *P* < 0.05 and |log_2_FC| > 2.0). Significant pathways (adjusted *P* < 0.05) for each cell type are listed in Table S3, and the top enriched pathways are shown in Figure 6. Astrocytic DEGs were enriched for *Parkinson’s Disease*, *Oxidative Phosphorylation*, *Huntington’s disease*, *Neurodegeneration*, and *Retrograde Endocannabinoid Signaling*. Endothelial DEGs were enriched for *Focal Adhesion*, *Integrin Signaling*, *Apoptosis*, *FoxO Signaling*, *PI3K-Akt Signaling*, and *Tight Junction*. Mature neuron DEGs were enriched for *Cocaine Addiction*, *Nicotine Addiction*, *Amphetamine Addiction*, *Neuroactive Ligand Signaling*, *MAPK Signaling*, and *Integrin Signaling*. Microglial DEGs were enriched for *Protein Processing in Endoplasmic Reticulum*, *Parathyroid Hormone Synthesis/Secretion/Action*, *Lysosome*, *Th17 Cell Differentiation*, *FoxO Signaling*, *Estrogen Signaling*, and *cGMP-PKG Signaling*. Oligodendrocytes DEGs were enriched for *Chemical Carcinogenesis (Reactive Oxygen Species)*, *Oxidative Phosphorylation*, *Parkinson’s Disease*, *Huntington’s Disease*, and *Estrogen Signaling*. OPC DEGs were enriched for *Steroid Biosynthesis* and *Terpenoid Backbone Biosynthesis*. Overall, DEGs in these six VTA cell types were significantly enriched for pathways related to mitochondrial function (oxidative phosphorylation), neurodegeneration (Parkinson’s disease, Huntington’s disease, neuroactive ligand-receptor interaction), and synaptic signaling (MAPK signaling). Notably, mature neuron DEGs were enriched for addiction-related pathways (cocaine, nicotine, amphetamine addiction), indicating neuron-specific transcriptional changes linked to substance-use mechanisms.

**Figure 6.**
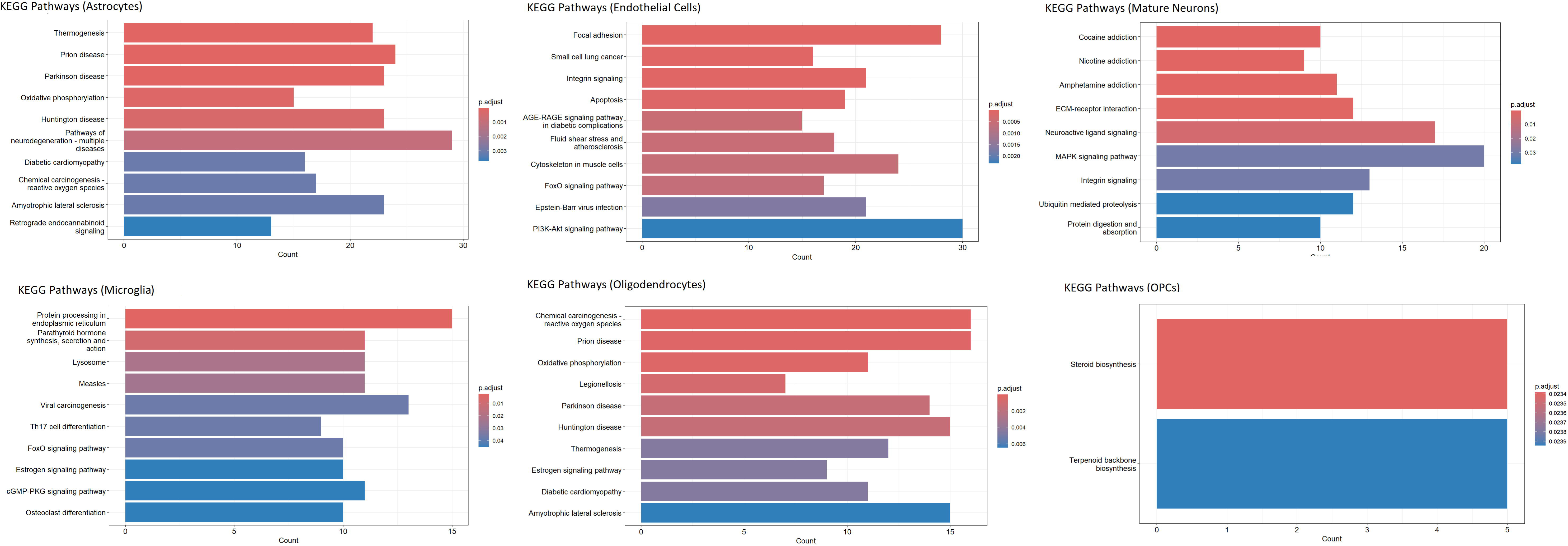
KEGG pathways enriched among DEGs in six VTA cell types.

### Functional enrichment of DEGs in four VTA neuronal subtypes

We performed KEGG pathway analysis on DEGs from four VTA neuronal subtypes (DEGs defined as *P* < 0.05 and |log_2_FC| > 2.0). Significant pathways (adjusted *P* < 0.05) for each neuronal subtypes are listed in Table S4, and the top enriched pathways are displayed in Figure 7. DEGs in dopaminergic neurons were overrepresented in KEGG pathways including *Synaptic Vesicle Cycle*, *Parkinson’s Disease*, *GABAergic Synapse*, *Folate Biosynthesis*, and *Neuroactive Ligand Signaling*. DEGs in GABAergic neurons were overrepresented in KEGG pathways including *Parkinson’ Disease*, *Chemical Carcinogenesis* (*Reactive Oxygen Species*), *Dopaminergic Synapse*, *Pathways of Neurodegeneration* (*Multiple Diseases*), *Huntington’s Disease*, *Alzheimer’s Disease*, *Oxidative Phosphorylation*, *Relaxin Signaling*, and *Retrograde Endocannabinoid Signaling*. DEGs in glutamatergic neurons were overrepresented in KEGG pathways including *Oxidative Phosphorylation*, *Parkinson’s Disease*, *Pathways of Neurodegeneration* (*Multiple Diseases*), *Apoptosis*, *Huntington’s Disease*, *Alzheimer’s Disease*, *Synaptic Vesicle Cycle*, *GABAergic Synapse*, and *Cocaine Addiction*. DEGs in unclassified neurons were overrepresented in KEGG pathways including *Amphetamine Addiction*, *Parathyroid Hormone Synthesis/Secretion/Action*, *Dopaminergic Synapse*, *Glutamatergic Synapse*, *Estrogen Signaling*, *Cocaine Addiction*, *Morphine Addiction*, *MAPK Signaling*, and *Cholinergic Synapse*. Collectively, neuronal DEGs highlight a convergence on mitochondrial/oxidative phosphorylation and neurodegeneration-related pathways across subtypes, while addiction- and synapse-related pathways show subtype-specific enrichment (notably dopaminergic and unclassified neurons), suggesting both shared and cell type-specific molecular perturbations in the VTA.

**Figure 7.**
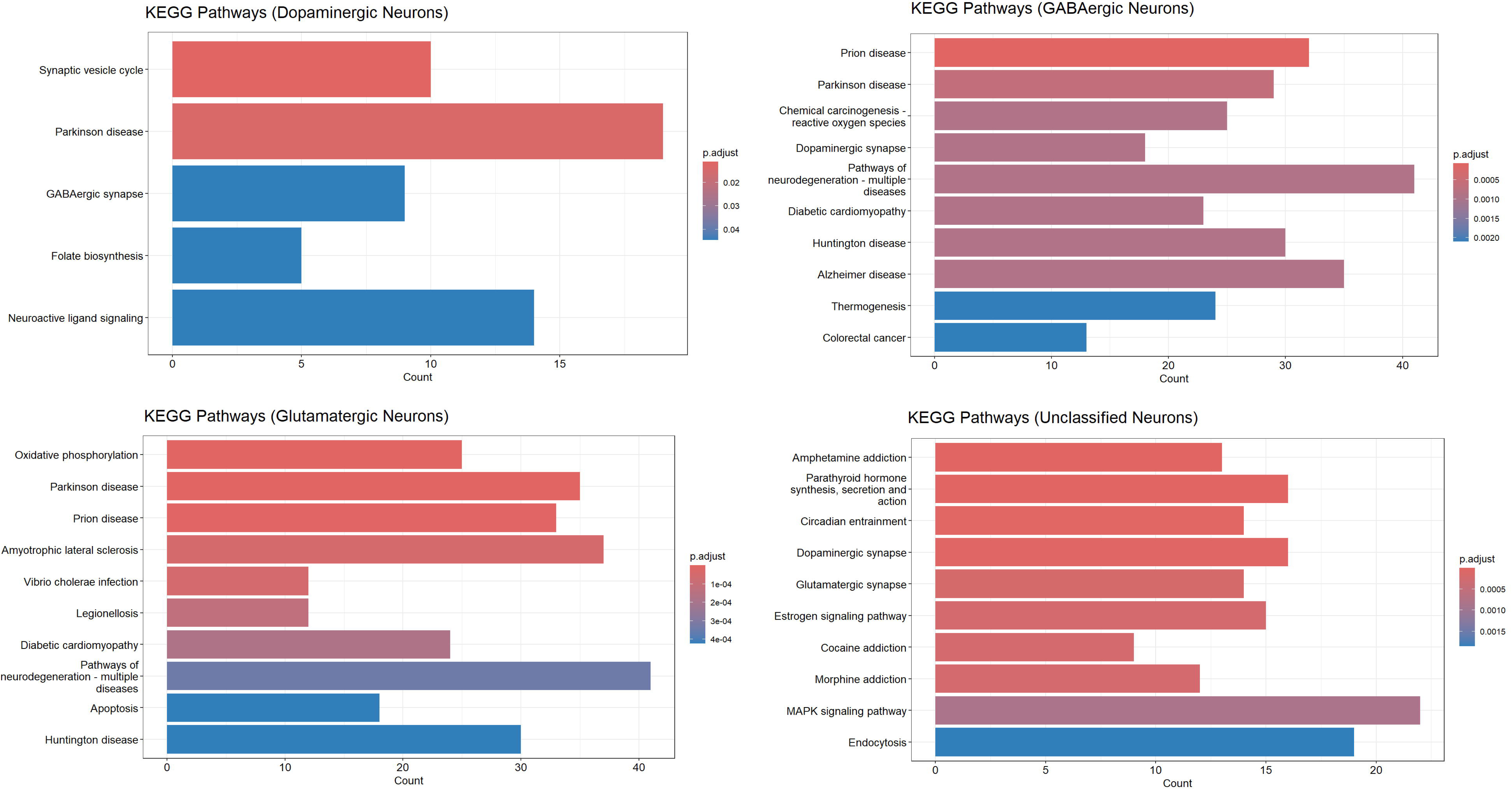
KEGG pathways enriched among DEGs in four VTA neuronal subtypes.

## CONCLUSIONS

### Identification of VTA cell type-specific transcriptomic alterations associated with AUD

Using snRNA-seq, we uncovered pronounced, cell type-specific transcriptomic dysregulation in the VTA of individuals with AUD. Clustering resolved six major cell types (astrocytes, endothelial cells, mature neurons, microglia, oligodendrocytes, and OPCs), and mature neurons were further subdivided into dopaminergic, GABAergic, glutamatergic, and unclassified subtypes. Across these cell populations, we detected hundreds of DEGs at *P* < 0.05 and |log_2_FC| > 2.0, with the largest DEG counts in endothelial cells, mature neurons, and GABAergic neurons. Two broad themes emerged: (1) widespread perturbation of mitochondrial/oxidative phosphorylation and neurodegeneration-related pathways across multiple cell types and neuronal subtypes, and (2) neuron subtype-specific enrichment of addiction- and synapse-related pathways, most prominently in dopaminergic and unclassified neurons. In addition, AUD-associated shifts in cellular composition (i.e., an increased proportion of endothelial cells and oligodendrocytes and decreased microglia) suggest cellular remodeling in the VTA. Together, these results provide a cell-resolved molecular portrait of AUD in a central reward region.

### Mature neuron and neuronal subtype-specific findings: functional and biological implications

Mature neurons showed extensive transcriptional dysregulation, and subdivision into neuronal subtypes highlighted both convergent and distinct responses to AUD. A shared signature across dopaminergic, GABAergic, and glutamatergic neurons was the enrichment of DEGs involved in mitochondrial function and oxidative phosphorylation, indicating that chronic alcohol exposure is associated with altered cellular energy metabolism and potential mitochondrial stress across neuronal classes. Mitochondrial dysfunction has been implicated in neurodegeneration and synaptic dysfunction (Akbar et al., 2016); in VTA neurons, compromised energy metabolism could impair action potential firing, neurotransmitter synthesis, synaptic vesicle cycling, and plasticity (Douma and de Kloet, 2020), all of which are critical for reward processing and addiction-related behavior.

Beyond this shared metabolic theme, dopaminergic neurons displayed a distinct enrichment for addiction-related and synaptic pathways (e.g., synaptic vesicle cycle, neuroactive ligand-receptor interactions), consistent with the central role of VTA dopamine neurons in reinforcement and alcohol-related reward (Juarez and Han, 2016). The predominance of upregulated DEGs in dopaminergic and glutamatergic neurons suggests compensatory or adaptive transcriptional responses, potentially reflecting homeostatic adjustments to repeated alcohol exposure or persistent alterations in signaling capacity. In contrast, the strong downregulation observed in GABAergic neurons may indicate impaired inhibitory control within the VTA microcircuitry, which could disinhibit dopaminergic neurons and contribute to aberrant reward signaling in AUD.

### Immediate-early genes and addiction-relevant transcriptional regulators

The differential expression of immediate-early genes (e.g., JUN, FOS/ΔFosB) across multiple cell types and neuronal subtypes aligns with known roles of these factors in activity-dependent plasticity and long-term adaptations to drugs of abuse (Rauscher et al., 1988). ΔFosB, in particular, has been implicated in the persistence of drug-induced behavioral changes (Nestler et al., 2001); its dysregulation across neuronal subtypes supports the notion of enduring transcriptional remodeling in the VTA of individuals with AUD.

### Non-neuronal cells and immune-related changes

Non-neuronal cells showed prominent, cell type-specific alterations that likely contribute to the VTA’s pathophysiology in AUD. Astrocytic DEGs were enriched for neurodegenerative and oxidative phosphorylation pathways, suggesting compromised metabolic and homeostatic support for neurons. Endothelial cells exhibited many upregulated DEGs and were enriched for pathways related to focal adhesion, tight junctions, PI3K-Akt, and FoxO signaling, indicative of possible vascular remodeling, blood-brain barrier (BBB) alterations, or endothelial stress in AUD. Such vascular changes could influence nutrient and oxygen delivery and modulate inflammatory signaling within the VTA. Microglia displayed fewer DEGs than some other glial types but were enriched for lysosomal and protein-processing pathways, consistent with altered phagocytic or immune-related activity (Hemati et al., 2025). Interestingly, we observed a reduced proportion of microglia in AUD cases relative to controls. Whether this reflects loss, phenotypic shift, or technical/nuclear-capture differences remains unclear, but it raises the possibility of altered neuroimmune surveillance or a dysregulated microglial response in chronic alcohol exposure. Oligodendrocyte and OPC DEGs implicate disruptions in myelination-related and lipid/sterol biosynthesis pathways, which could affect axonal conduction and VTA connectivity.

### Shared molecular signatures and potential mechanistic links

The convergence on mitochondrial/oxidative phosphorylation and pathways typically associated with neurodegenerative diseases (Parkinson’s, Huntington’s, Alzheimer’s) across cell types suggests that chronic alcohol exposure induces stress responses that overlap with general neurodegenerative processes. Heat shock protein genes (*HSPA1A*, *HSPA1B*, *HSPB1*) being differentially expressed across multiple cell types supports broad cellular stress and protein-homeostasis dysregulation. Additionally, we observed downregulation of *ALDH2* in endothelial cells, oligodendrocytes, and OPCs. *ALDH2* encodes a mitochondrial aldehyde dehydrogenase that metabolizes acetaldehyde and other reactive aldehydes (Vasiliou and Nebert, 2005); its dysregulation in specific VTA cell types suggests altered local aldehyde detoxification and redox balance. In endothelial cells, reduced ALDH2 could impair blood-brain barrier (BBB) integrity and neurovascular function, increasing permeability, promoting inflammatory signaling, and enhancing exposure of the parenchyma to peripheral inflammatory mediators or ethanol metabolites. In oligodendrocytes and OPCs, diminished ALDH2 may increase vulnerability of myelin lipids and proteins to aldehyde-mediated damage, with potential consequences for myelin maintenance, white-matter integrity, and network connectivity. Together, these changes could contribute indirectly to oxidative stress, neuroinflammation, and the connectivity and cognitive deficits reported in AUD.

### Comparison with prior studies

Our cell-resolved results extend prior bulk and region-level transcriptomic studies of AUD by assigning molecular changes to discrete VTA cell populations. Consistent with bulk reports of mitochondrial and synaptic pathway dysregulation in AUD-affected brain regions (Arzua et al., 2024), we observed similar themes but reveal that these processes are distributed across cell types with subtype-specific nuances. Moreover, compared with our previous human postmortem VTA bulk RNA seq-AUD results (Besong et al., 2024), gene level concordance was limited, but pathway level overlap was observed: *Neurodegeneration*, *Apoptosis*, and *Adherens Junction* pathways were identified in both bulk and single nucleus datasets. The dopaminergic neuron-specific enrichment for addiction-related pathways corroborates animal-model work implicating VTA dopamine circuits in alcohol reinforcement (Morel et al., 2019) and supports the translational relevance of preclinical findings to human AUD.

### Potential clinical and translational implications

The cell type-specific DEGs and pathways identified here nominate candidate molecular targets for therapeutic intervention. For instance, strategies aimed at preserving mitochondrial function or reducing oxidative stress in susceptible VTA neurons and glia might mitigate alcohol-induced dysfunction. Modulating synaptic vesicle cycling or receptor signaling selectively in dopaminergic neurons could also normalize maladaptive reward signaling. Endothelial and BBB-targeted interventions could influence neuroinflammation and metabolic support. Importantly, several DEGs (including immediate-early genes and heat shock proteins) are tractable to pharmacologic modulation or can be evaluated as potential biomarkers of VTA dysfunction in AUD.

### Limitations

Several limitations temper interpretation. First and foremost, the sample size is small. The present study includes only one biological replicate per sex/group. Consequently, individual-specific variation cannot be excluded, and results should therefore be considered exploratory and hypothesis-generating rather than definitive. Future work with larger, adequately powered cohorts is planned to validate these findings. Second, snRNA-seq captures nuclear and nascent transcripts and may not fully reflect cytoplasmic mRNA dynamics or protein-level changes; follow-up at the protein level and with spatially resolved approaches would strengthen the biological interpretation. Third, although cases and controls were matched on basic factors and screened for major confounds, residual heterogeneity in lifetime drinking patterns, comorbid conditions, medication, smoking, PMI, and agonal state could influence expression. Fourth, the cross-sectional, postmortem design precludes causal inference. The observed transcriptional differences may reflect consequences of chronic alcohol exposure, predisposing factors, or terminal events. Fifth, some inferred cell composition differences may be influenced by technical factors inherent in nuclei isolation and capture or by differential nuclear RNA content across cell types. Finally, we used unadjusted *P* < 0.05 and |log2FC| > 2.0 thresholds for DEG calling in this small cohort; while these criteria helped highlight robust changes, they may also include false positives and should be validated in larger cohorts. Key next steps include replication in larger, independent postmortem VTA cohorts with richer clinical metadata (e.g., quantity/duration of alcohol use, last drink, comorbidities, medication history) to assess reproducibility and to enable stratified analyses (sex, severity, comorbidity). Integration with spatial transcriptomics would map the anatomical distribution of cell type-specific changes within VTA microcircuits. Multi-omic approaches (e.g., proteomics, epigenomics, and metabolomics) could clarify whether transcript-level changes translate to functional protein and metabolic alterations. Functional studies using human cellular models (iPSC-derived midbrain neurons and glia) or targeted animal experiments could test causal roles for high-priority DEGs (e.g., JUN, FOS/ΔFosB, ALDH2, HSPs) and evaluate therapeutic modulation.

### Summary

This snRNA-seq study provides the first cell type-resolved transcriptomic atlas of the human VTA in AUD, revealing widespread and cell-specific molecular perturbations. Our data highlight convergent mitochondrial and neurodegeneration-related alterations across cell types alongside neuron subtype-specific changes in addiction- and synapse-related pathways, particularly within dopaminergic neurons. Although limited by sample size, these findings generate testable hypotheses about cell-resolved mechanisms of AUD in the human VTA and identify candidate targets for mechanistic follow-up and potential therapeutic development.

## ACKNOWLEDGMENTS

This work was supported by a Boston University Genome Sciences Institute (GSI) Pilot Grant, the National Institute on Alcohol Abuse and Alcoholism (NIAAA) grant R01AA029758, and the National Institute of Diabetes and Digestive and Kidney Diseases (NIDDK) grant R21DK143406. We thank Yuriy Alekseyev, Christopher Williams, and Salam Alabdullatif from the Boston University Medical Campus Single Cell Sequencing Core for their assistance. We are also grateful to the New South Wales Brain Tissue Resource Centre (NSWBTRC) at the University of Sydney for providing alcoholic and control brain tissues for this study. The NSWBTRC is supported by the University of Sydney, the National Health and Medical Research Council of Australia, and the NIAAA. Finally, we thank the deceased donors’ next of kin for providing informed written consent for tissue donation and research use.

## DATA AVAILABILITY STATEMENT

All data produced in the present study are available upon reasonable request to the authors.

## REFERENCES

1. Akbar M, Essa MM, Daradkeh G, Abdelmegeed MA, Choi Y, Mahmood L, Song BJ (2016) Mitochondrial dysfunction and cell death in neurodegenerative diseases through nitroxidative stress. Brain research 1637:34–55.

2. American Psychiatric Association (1994) American Psychiatric Association: Diagnostic and Statistical Manual of Mental Disorders, 4th ed. Washington, DC, American Psychiatric Association.

3. Arzua T, Yan Y, Liu X, Dash RK, Liu QS, Bai X (2024) Synaptic and mitochondrial mechanisms behind alcohol-induced imbalance of excitatory/inhibitory synaptic activity and associated cognitive and behavioral abnormalities. Translational psychiatry 14:51.

4. Besong OTO, Koo JS, Zhang H (2024) Brain lncRNA-mRNA co-expression regulatory networks and alcohol use disorder. Genomics 116:110928.

5. Blighe K, Rana S, Lewis M (2025) EnhancedVolcano: Publication-ready volcano plots with enhanced colouring and labeling. R package version 1291, https://bioconductororg/packages/EnhancedVolcano/.

6. Brenner E, Tiwari GR, Kapoor M, Liu Y, Brock A, Mayfield RD (2020) Single cell transcriptome profiling of the human alcohol-dependent brain. Hum Mol Genet 29:1144–1153.

7. Douma EH, de Kloet ER (2020) Stress-induced plasticity and functioning of ventral tegmental dopamine neurons. Neurosci Biobehav Rev 108:48–77.

8. Farris SP, Arasappan D, Hunicke-Smith S, Harris RA, Mayfield RD (2015) Transcriptome organization for chronic alcohol abuse in human brain. Molecular psychiatry 20:1438–1447.

9. Flatscher-Bader T, van der Brug M, Hwang JW, Gochee PA, Matsumoto I, Niwa S, Wilce PA (2005) Alcohol-responsive genes in the frontal cortex and nucleus accumbens of human alcoholics. J Neurochem 93:359–370.

10. Grindberg RV, Yee-Greenbaum JL, McConnell MJ, Novotny M, O’Shaughnessy AL, Lambert GM, Arauzo-Bravo MJ, Lee J, Fishman M, Robbins GE, Lin X, Venepally P, Badger JH, Galbraith DW, Gage FH, Lasken RS (2013) RNA-sequencing from single nuclei. Proceedings of the National Academy of Sciences of the United States of America 110:19802–19807.

11. Hao Y, Stuart T, Kowalski MH, Choudhary S, Hoffman P, Hartman A, Srivastava A, Molla G, Madad S, Fernandez-Granda C, Satija R (2024) Dictionary learning for integrative, multimodal and scalable single-cell analysis. Nature biotechnology 42:293–304.

12. Hemati H, Blanton MB, True HE, Koura J, Khadka R, Grant KA, Messaoudi I (2025) Chronic alcohol consumption induces phenotypic and functional alterations consistent with a hyper-inflammatory state in peripheral blood mononuclear cell-derived microglia in a rhesus macaque model. Brain Behav Immun 129:874–889.

13. Ianevski A, Giri AK, Aittokallio T (2022) Fully-automated and ultra-fast cell-type identification using specific marker combinations from single-cell transcriptomic data. Nature communications 13:1246.

14. Juarez B, Han MH (2016) Diversity of Dopaminergic Neural Circuits in Response to Drug Exposure. Neuropsychopharmacology 41:2424–2446.

15. Koob GF (2008) A role for brain stress systems in addiction. Neuron 59:11–34.

16. Koob GF, Volkow ND (2016) Neurobiology of addiction: a neurocircuitry analysis. Lancet Psychiatry 3:760–773.

17. Korsunsky I, Millard N, Fan J, Slowikowski K, Zhang F, Wei K, Baglaenko Y, Brenner M, Loh PR, Raychaudhuri S (2019) Fast, sensitive and accurate integration of single-cell data with Harmony. Nat Methods 16:1289–1296.

18. Lake BB, Ai R, Kaeser GE, Salathia NS, Yung YC, Liu R, Wildberg A, Gao D, Fung HL, Chen S, Vijayaraghavan R, Wong J, Chen A, Sheng X, Kaper F, Shen R, Ronaghi M, Fan JB, Wang W, Chun J, Zhang K (2016) Neuronal subtypes and diversity revealed by single-nucleus RNA sequencing of the human brain. Science 352:1586–1590.

19. Lewohl JM, Wang L, Miles MF, Zhang L, Dodd PR, Harris RA (2000) Gene expression in human alcoholism: microarray analysis of frontal cortex. Alcohol Clin Exp Res 24:1873–1882.

20. Lex A, Gehlenborg N, Strobelt H, Vuillemot R, Pfister H (2014) UpSet: Visualization of Intersecting Sets. IEEE Trans Vis Comput Graph 20:1983–1992.

21. Lim Y, Beane-Ebel JE, Tanaka Y, Ning B, Husted CR, Henderson DC, Xiang Y, Park IH, Farrer LA, Zhang H (2021) Exploration of alcohol use disorder-associated brain miRNA-mRNA regulatory networks. Translational psychiatry 11:504.

22. Liu J, Lewohl JM, Dodd PR, Randall PK, Harris RA, Mayfield RD (2004) Gene expression profiling of individual cases reveals consistent transcriptional changes in alcoholic human brain. J Neurochem 90:1050–1058.

23. Liu J, Lewohl JM, Harris RA, Iyer VR, Dodd PR, Randall PK, Mayfield RD (2006) Patterns of gene expression in the frontal cortex discriminate alcoholic from nonalcoholic individuals. Neuropsychopharmacology 31:1574–1582.

24. Mayfield J, Ferguson L, Harris RA (2013) Neuroimmune signaling: a key component of alcohol abuse. Current opinion in neurobiology 23:513–520.

25. Mayfield RD, Lewohl JM, Dodd PR, Herlihy A, Liu J, Harris RA (2002) Patterns of gene expression are altered in the frontal and motor cortices of human alcoholics. J Neurochem 81:802–813.

26. McClintick JN, Xuei X, Tischfield JA, Goate A, Foroud T, Wetherill L, Ehringer MA, Edenberg HJ (2013) Stress-response pathways are altered in the hippocampus of chronic alcoholics. Alcohol 47:505–515.

27. McGinnis CS, Murrow LM, Gartner ZJ (2019) DoubletFinder: Doublet Detection in Single-Cell RNA Sequencing Data Using Artificial Nearest Neighbors. Cell Syst 8:329–337 e324.

28. McReynolds JR, Christianson JP, Blacktop JM, Mantsch JR (2018) What does the Fos say? Using Fos-based approaches to understand the contribution of stress to substance use disorders. Neurobiol Stress 9:271–285.

29. Morel C, Montgomery S, Han MH (2019) Nicotine and alcohol: the role of midbrain dopaminergic neurons in drug reinforcement. Eur J Neurosci 50:2180–2200.

30. Nader K, Tasci M, Ianevski A, Erickson A, Verschuren EW, Aittokallio T, Miihkinen M (2024) ScType enables fast and accurate cell type identification from spatial transcriptomics data. Bioinformatics 40:btae426.

31. Nestler EJ, Barrot M, Self DW (2001) DeltaFosB: a sustained molecular switch for addiction. Proceedings of the National Academy of Sciences of the United States of America 98:11042–11046.

32. Ponomarev I, Wang S, Zhang L, Harris RA, Mayfield RD (2012) Gene coexpression networks in human brain identify epigenetic modifications in alcohol dependence. J Neurosci 32:1884–1897.

33. Rauscher FJ, 3rd, Voulalas PJ, Franza BR, Jr., Curran T (1988) Fos and Jun bind cooperatively to the AP-1 site: reconstitution in vitro. Genes Dev 2:1687–1699.

34. van den Oord E, Xie LY, Zhao M, Aberg KA, Clark SL (2023) A single-nucleus transcriptomics study of alcohol use disorder in the nucleus accumbens. Addict Biol 28:e13250.

35. Vasiliou V, Nebert DW (2005) Analysis and update of the human aldehyde dehydrogenase (ALDH) gene family. Human genomics 2:138–143.

36. Yao Z, van Velthoven CTJ, Nguyen TN, Goldy J, Sedeno-Cortes AE, Baftizadeh F, Bertagnolli D, Casper T, Chiang M, Crichton K, Ding SL, Fong O, Garren E, Glandon A, Gouwens NW, Gray J, Graybuck LT, Hawrylycz MJ, Hirschstein D, Kroll M, Lathia K, Lee C, Levi B, McMillen D, Mok S, Pham T, Ren Q, Rimorin C, Shapovalova N, Sulc J, Sunkin SM, Tieu M, Torkelson A, Tung H, Ward K, Dee N, Smith KA, Tasic B, Zeng H (2021) A taxonomy of transcriptomic cell types across the isocortex and hippocampal formation. Cell 184:3222–3241 e3226.

37. Yu G, Wang LG, Han Y, He QY (2012) clusterProfiler: an R package for comparing biological themes among gene clusters. OMICS 16:284–287.

38. Zhang H, Wang F, Xu H, Liu Y, Liu J, Zhao H, Gelernter J (2014) Differentially co-expressed genes in postmortem prefrontal cortex of individuals with alcohol use disorders: influence on alcohol metabolism-related pathways. Hum Genet 133:1383–1394.

39. Zheng GX, Terry JM, Belgrader P, Ryvkin P, Bent ZW, Wilson R, Ziraldo SB, Wheeler TD, McDermott GP, Zhu J, Gregory MT, Shuga J, Montesclaros L, Underwood JG, Masquelier DA, Nishimura SY, Schnall-Levin M, Wyatt PW, Hindson CM, Bharadwaj R, Wong A, Ness KD, Beppu LW, Deeg HJ, McFarland C, Loeb KR, Valente WJ, Ericson NG, Stevens EA, Radich JP, Mikkelsen TS, Hindson BJ, Bielas JH (2017) Massively parallel digital transcriptional profiling of single cells. Nature communications 8:14049.

40. Zhou Z, Enoch MA, Goldman D (2014) Gene expression in the addicted brain. International review of neurobiology 116:251–273.

