## Supplemental Materials for "Single-nucleus RNA sequencing identifies transcriptomic signatures of alcohol use disorder in the human ventral tegmental area"

(Supporting Information)

Figure S1. Electropherogram of the constructed sequencing library analyzed by Agilent Bioanalyzer.

Figure S2 The experimental workflow.

Figure S3 Quality control metrics (detected features, counts, and percentages of mitochondrial and ribosomal RNA).

Figure S4 Doublets predicted with DoubletFinder.

Figure S5 Unintegrated and harmony clustering analyses of all samples, as well as analyses stratified by sample identity and subject gender.

Table S1 Details of four human postmortem ventral tegmental area (VTA) tissue samples.

Table S2 Quality control metrics for extracted nuclei and snRNA-Seq data.

Table S3 KEGG pathways enriched among DEGs in six VTA cell types.

Table S4 KEGG pathways enriched among DEGs in four VTA neuronal subtypes.

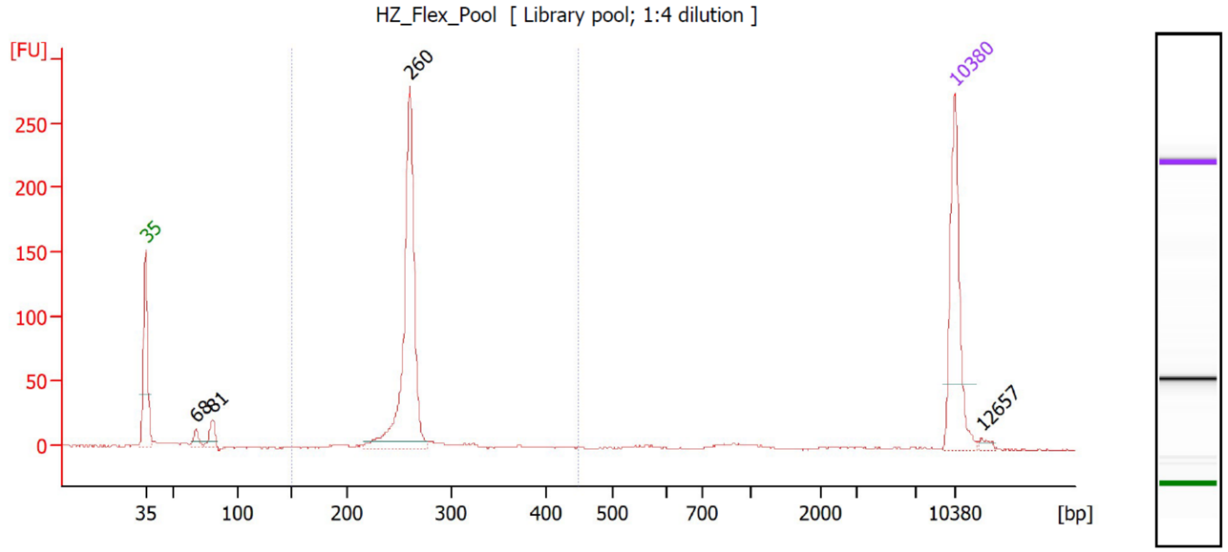

Figure S1. Electropherogram of the constructed sequencing library analyzed by Agilent Bioanalyzer.

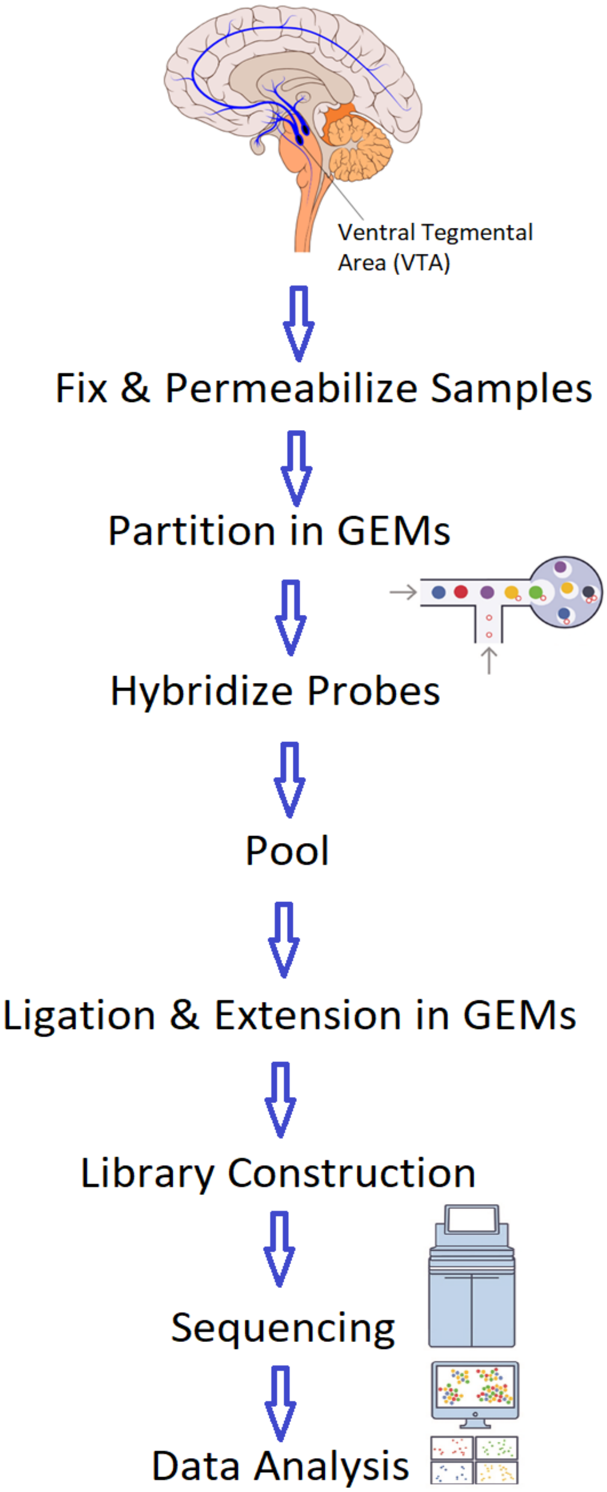

Figure S2 The experimental workflow.

**Fig. S3a**

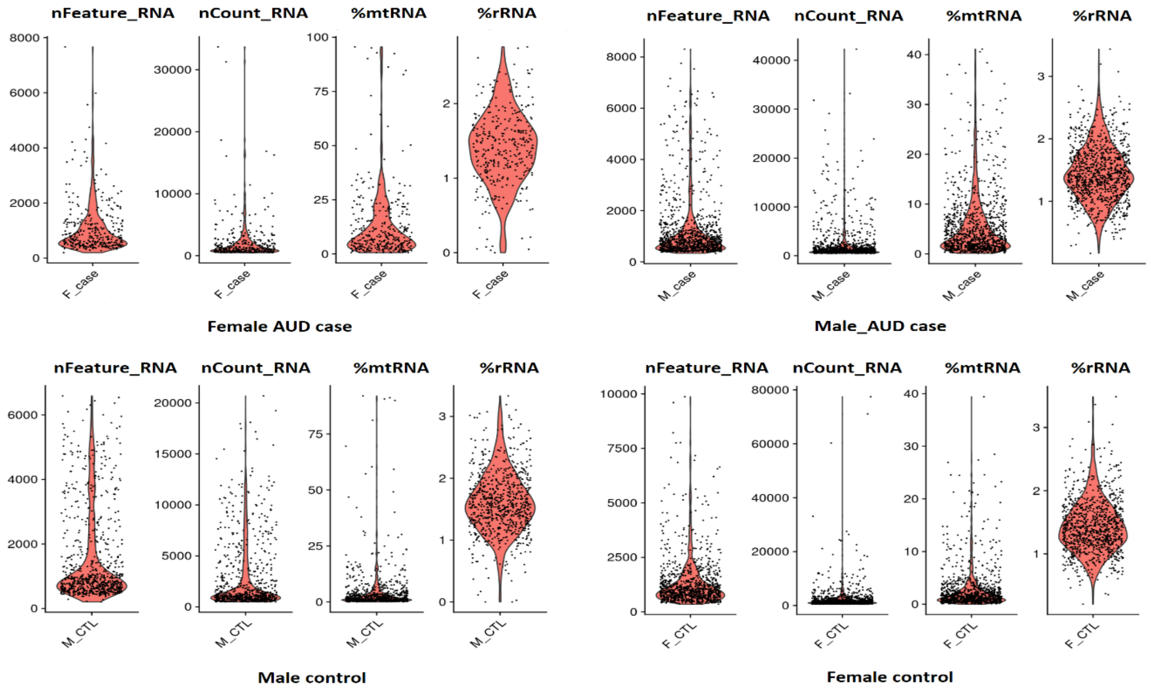

**Fig. S3b**

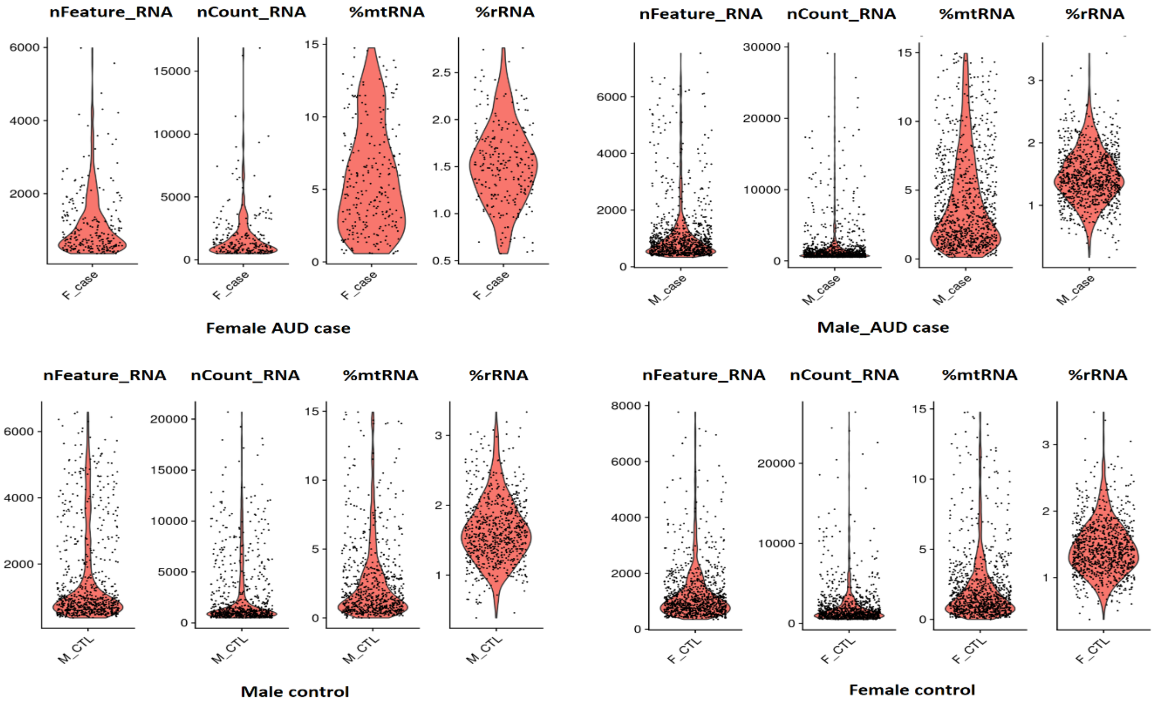

Figure S3. Quality control metrics (detected features, counts, and percentages of mitochondrial and ribosomal RNA).
Fig. S3a. Quality control metrics before filtration; Fig. S3b. Quality control metrics after filtration.

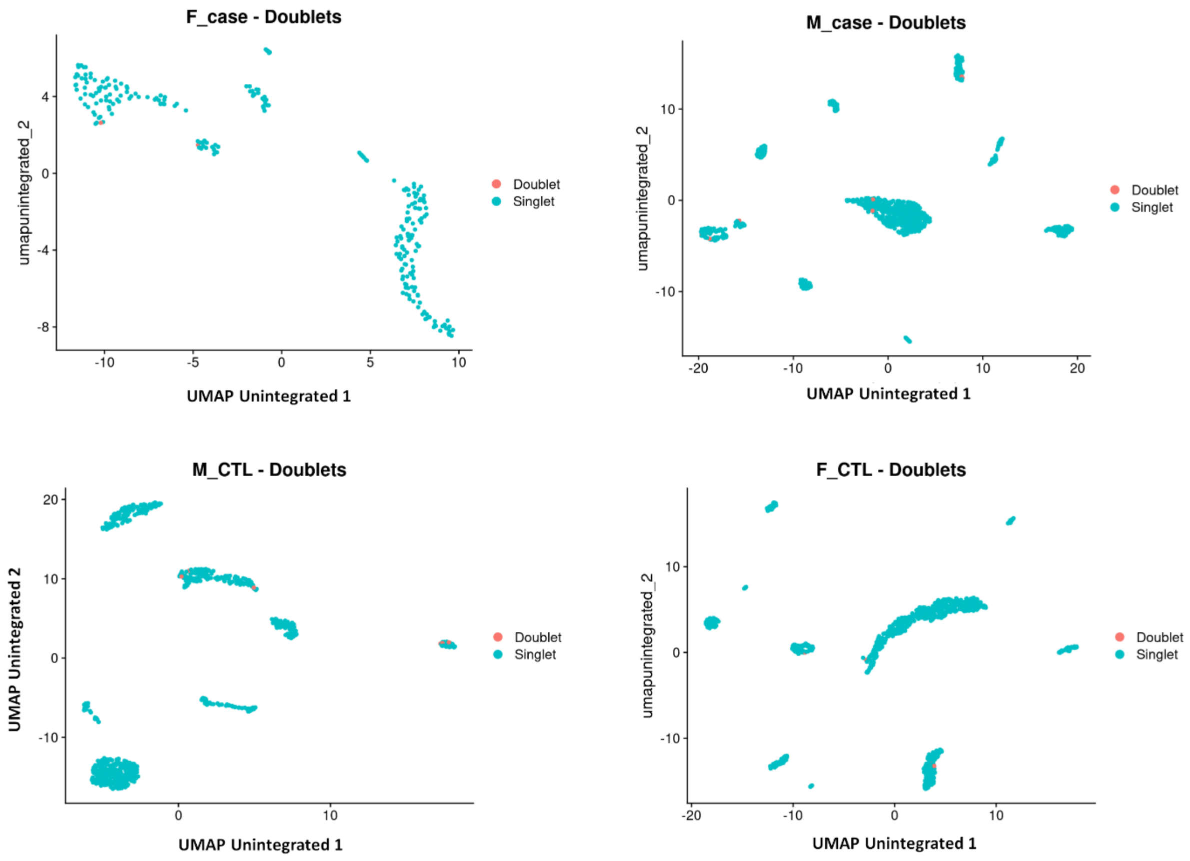

Figure S4 Doublets predicted with DoubletFinder.
Very few doublets were detected in the four samples (female AUD case, male AUD case, male control, and female control)

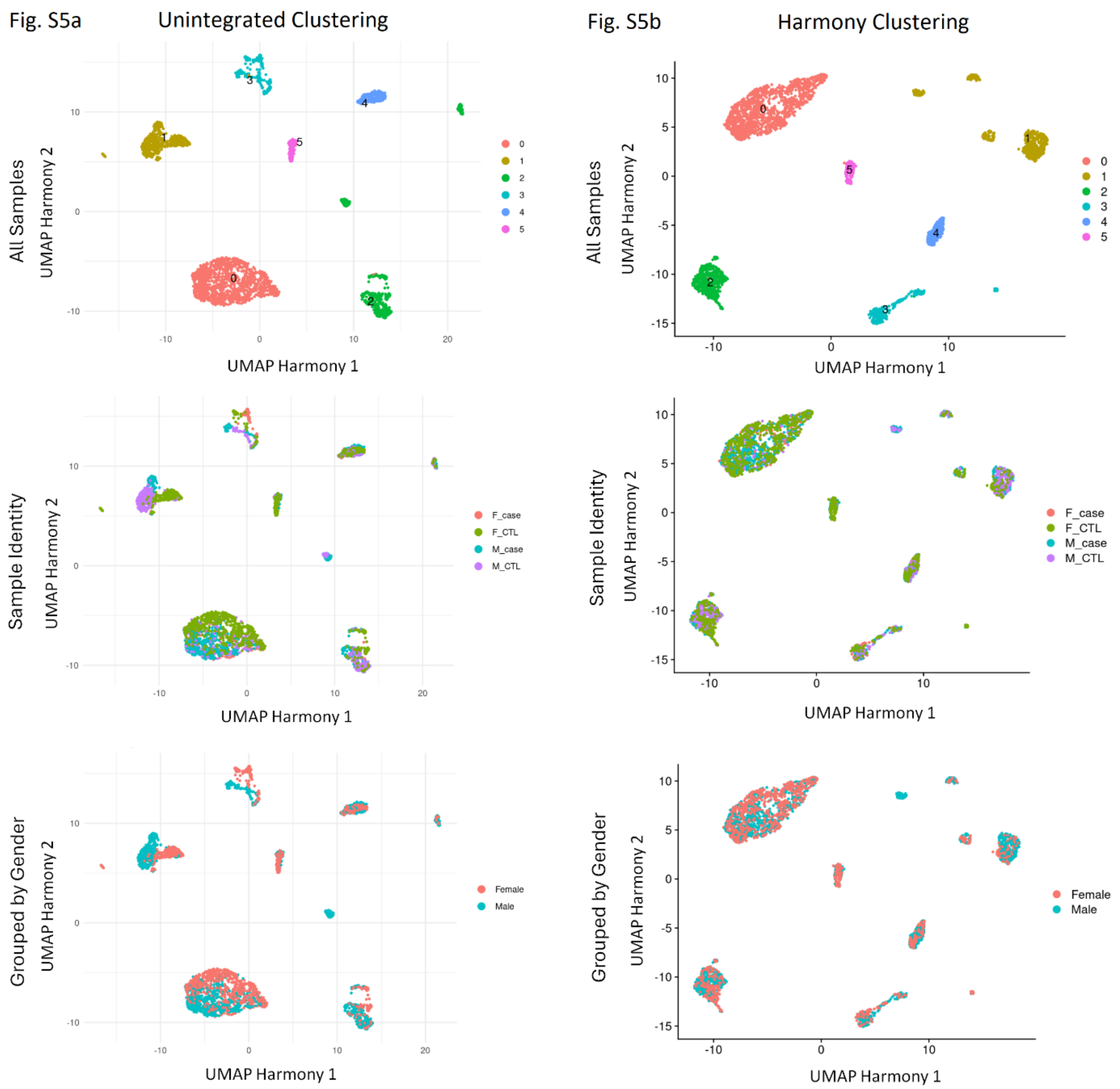

Figure S5 Unintegrated and harmony clustering analyses of all samples, as well as analyses stratified by sample identity and subject gender.
Fig. S5a: Unintegrated clustering analysis; Fig. S5b: Harmony clustering analysis

SUPPLEMENTAL TABLE 1 Details of four human postmortem ventral tegmental area (VTA) tissue samples.

| Sample ID | HZ_1 | HZ_2 | HZ_3 | HZ_4 |
|---|---|---|---|---|
| Ethnic Origin | European | European | European | European |
| Classification | Alcohol use disorder | Alcohol use disorder | Control | Control |
| Sex | Female | Male | Male | Female |
| Age | 67 | 58 | 62 | 74 |
| Brain Weight (gram) | 1,308 | 1,250 | 1,430 | 1,184 |
| Postmortem Interval (PMI) (hours) | 18 | 44.5 | 46 | 20 |
| Brain tissue pH | 5.89 | 6.47 | 6.95 | 6.59 |
| Cause of death category | Hepatic | Cardiac | Cardiac | Cancer |
| Alcohol Intake(g/day) | 135 | 110 | 2 | 20 |

SUPPLEMENTAL TABLE 2 Quality control metrics for extracted nuclei and snRNA-Seq data.

| Sample ID | HZ_1 | HZ_2 | HZ_3 | HZ_4 |
|---|---|---|---|---|
| Number of nuclei/ml | 2,220,000 | 2,450,000 | 2,280,000 | 1,585,000 |
| Viability of nuclei | 80.0% | 75.0% | 98.0% | 87.0% |
| Mean reads per nucleus | 36,101 | 37,963 | 55,809 | 43,478 |
| Total reads per sample | 35,522,980 | 51,060,164 | 46,321,184 | 56,434,454 |
| Estimated number of nuclei captured | 984 | 1,345 | 830 | 1,298 |
| Probe barcode ID | BC001 | BC002 | BC003 | BC004 |
| UMIs per probe barcode | 1,631,834 (12.8%) | 3,842,521 (30.1%) | 3,336,124 (26.1%) | 3,940,926 (30.9%) |
| Cells per probe barcode | 343 (11%) | 1,024 (32.8%) | 770 (24.7%) | 988 (31.6%) |
| Number of nuclei before QC filtering | 343 | 1024 | 770 | 988 |
| Number of detected genes before QC filtering | 12556 | 14231 | 14293 | 14264 |
| Number of nuclei after QC filtering | 249 | 945 | 721 | 959 |
| Number of detected genes after QC filtering | 12556 | 14231 | 14293 | 14264 |

SUPPLEMENTAL TABLE 3 KEGG pathways enriched among DEGs in six VTA cell types.

| KEGG Pathways | GeneRatio | Fold Enrichment | *P*-value | *P*adj |
|---|---|---|---|---|
| **Astrocytes** | | | | |
| Thermogenesis | 22/248 | 3.59 | 1.87E-07 | 3.25E-05 |
| Prion disease | 24/248 | 3.31 | 2.33E-07 | 3.25E-05 |
| Parkinson disease | 23/248 | 3.26 | 5.70E-07 | 5.30E-05 |
| Oxidative phosphorylation | 15/248 | 4.17 | 2.83E-06 | 0.00020 |
| Huntington disease | 23/248 | 2.84 | 6.06E-06 | 0.00034 |
| Pathways of neurodegeneration - multiple diseases | 29/248 | 2.31 | 2.15E-05 | 0.0010 |
| Diabetic cardiomyopathy | 16/248 | 3.00 | 8.75E-05 | 0.0031 |
| Chemical carcinogenesis - reactive oxygen species | 17/248 | 2.88 | 8.81E-05 | 0.0031 |
| Amyotrophic lateral sclerosis | 23/248 | 2.38 | 9.94E-05 | 0.0031 |
| Retrograde endocannabinoid signaling | 13/248 | 3.35 | 0.00013 | 0.0037 |
| SNARE interactions in vesicular transport | 6/248 | 6.98 | 0.00018 | 0.0046 |
| Lysine degradation | 8/248 | 4.88 | 0.00021 | 0.0049 |
| **Endothelial Cells** | | | | |
| Focal adhesion | 28/371 | 3.54 | 4.67E-09 | 1.46E-06 |
| Small cell lung cancer | 16/371 | 4.42 | 5.01E-07 | 5.27E-05 |
| Integrin signaling | 21/371 | 3.50 | 5.05E-07 | 5.27E-05 |
| Apoptosis | 19/371 | 3.56 | 1.37E-06 | 0.00011 |
| AGE-RAGE signaling pathway in diabetic complications | 15/371 | 3.81 | 7.64E-06 | 0.00048 |
| Fluid shear stress and atherosclerosis | 18/371 | 3.25 | 9.48E-06 | 0.00049 |
| Cytoskeleton in muscle cells | 24/371 | 2.64 | 1.26E-05 | 0.00056 |
| FoxO signaling pathway | 17/371 | 3.28 | 1.50E-05 | 0.00059 |
| Epstein-Barr virus infection | 21/371 | 2.63 | 4.78E-05 | 0.0017 |
| PI3K-Akt signaling pathway | 30/371 | 2.13 | 7.44E-05 | 0.0023 |
| Human papillomavirus infection | 28/371 | 2.16 | 0.00010 | 0.0028 |
| Tight junction | 18/371 | 2.72 | 0.00011 | 0.0028 |
| Proteoglycans in cancer | 20/371 | 2.52 | 0.00013 | 0.0032 |
| Bladder cancer | 8/371 | 5.01 | 0.00015 | 0.0034 |
| ECM-receptor interaction | 12/371 | 3.46 | 0.00016 | 0.0034 |
| Chronic myeloid leukemia | 11/371 | 3.67 | 0.00018 | 0.0035 |
| Regulation of actin cytoskeleton | 21/371 | 2.32 | 0.00028 | 0.0051 |
| Platelet activation | 14/371 | 2.85 | 0.00038 | 0.0066 |
| Endometrial cancer | 9/371 | 3.91 | 0.00042 | 0.0069 |
| Non-small cell lung cancer | 10/371 | 3.52 | 0.00050 | 0.0078 |
| Arginine and proline metabolism | 8/371 | 4.11 | 0.00063 | 0.0094 |
| Glioma | 10/371 | 3.38 | 0.00069 | 0.0098 |
| JAK-STAT signaling pathway | 16/371 | 2.44 | 0.00084 | 0.011 |
| Shigellosis | 21/371 | 2.13 | 0.00088 | 0.011 |
| Cellular senescence | 15/371 | 2.45 | 0.0012 | 0.015 |
| Prion disease | 22/371 | 2.03 | 0.0012 | 0.015 |

SUPPLEMENTAL TABLE 3 (cont.)

| | | | | |
|---|---|---|---|---|
| MAPK signaling pathway | 23/371 | 1.97 | 0.0015 | 0.017 |
| Fc gamma R-mediated phagocytosis | 11/371 | 2.85 | 0.0016 | 0.018 |
| Central carbon metabolism in cancer | 9/371 | 3.25 | 0.0017 | 0.018 |
| Breast cancer | 14/371 | 2.43 | 0.0019 | 0.020 |
| Colorectal cancer | 10/371 | 2.95 | 0.0020 | 0.020 |
| Melanoma | 9/371 | 3.16 | 0.0020 | 0.020 |
| Parkinson disease | 21/371 | 1.99 | 0.0021 | 0.020 |
| Prostate cancer | 11/371 | 2.66 | 0.0027 | 0.025 |
| Thyroid cancer | 6/371 | 4.16 | 0.0028 | 0.025 |
| Pancreatic cancer | 9/371 | 3.00 | 0.0029 | 0.025 |
| Influenza A | 15/371 | 2.23 | 0.0031 | 0.026 |
| Bacterial invasion of epithelial cells | 9/371 | 2.96 | 0.0032 | 0.026 |
| IgSF CAM signaling | 22/371 | 1.87 | 0.0035 | 0.028 |
| Salmonella infection | 19/371 | 1.94 | 0.0043 | 0.034 |
| Endocytosis | 19/371 | 1.93 | 0.0045 | 0.034 |
| Cobalamin transport and metabolism | 4/371 | 5.70 | 0.0045 | 0.034 |
| Acute myeloid leukemia | 8/371 | 3.02 | 0.0047 | 0.034 |
| ErbB signaling pathway | 9/371 | 2.69 | 0.0061 | 0.044 |
| Amoebiasis | 10/371 | 2.49 | 0.0067 | 0.047 |
| **Microglial Cells** | | | | |
| Protein processing in endoplasmic reticulum | 15/219 | 3.79 | 9.45E-06 | 0.0026 |
| Parathyroid hormone synthesis, secretion and action | 11/219 | 4.16 | 6.48E-05 | 0.0087 |
| Lysosome | 11/219 | 3.60 | 0.00024 | 0.022 |
| Measles | 11/219 | 3.44 | 0.00035 | 0.024 |
| Viral carcinogenesis | 13/219 | 2.76 | 0.00089 | 0.037 |
| Th17 cell differentiation | 9/219 | 3.59 | 0.00089 | 0.037 |
| FoxO signaling pathway | 10/219 | 3.27 | 0.00097 | 0.037 |
| Estrogen signaling pathway | 10/219 | 3.13 | 0.0014 | 0.045 |
| cGMP-PKG signaling pathway | 11/219 | 2.88 | 0.0015 | 0.045 |
| Osteoclast differentiation | 10/219 | 3.04 | 0.0017 | 0.045 |
| **Mature Neurons** | | | | |
| Cocaine addiction | 10/285 | 6.82 | 1.44E-06 | 0.00038 |
| Nicotine addiction | 9/285 | 7.33 | 2.57E-06 | 0.00038 |
| Amphetamine addiction | 11/285 | 5.33 | 5.55E-06 | 0.00055 |
| ECM-receptor interaction | 12/285 | 4.50 | 1.24E-05 | 0.00092 |
| Neuroactive ligand signaling | 17/285 | 2.85 | 9.57E-05 | 0.0057 |
| MAPK signaling pathway | 20/285 | 2.23 | 0.00067 | 0.0300 |
| Integrin signaling | 13/285 | 2.82 | 0.00071 | 0.0300 |
| Ubiquitin mediated proteolysis | 12/285 | 2.82 | 0.0011 | 0.0382 |
| Protein digestion and absorption | 10/285 | 3.18 | 0.0012 | 0.0382 |

SUPPLEMENTAL TABLE 3 (cont.)

| | | | | |
|---|---|---|---|---|
| **Oliogodendrocytes** | | | | |
| Chemical carcinogenesis - reactive oxygen species | 16/137 | 4.90 | 1.47E-07 | 3.42E-05 |
| Prion disease | 16/137 | 4.00 | 2.24E-06 | 0.00026 |
| Oxidative phosphorylation | 11/137 | 5.54 | 4.44E-06 | 0.00034 |
| Legionellosis | 7/137 | 8.69 | 1.41E-05 | 0.00082 |
| Parkinson disease | 14/137 | 3.59 | 3.38E-05 | 0.0015 |
| Huntington disease | 15/137 | 3.35 | 3.89E-05 | 0.0015 |
| Thermogenesis | 12/137 | 3.55 | 0.00014 | 0.0045 |
| Estrogen signaling pathway | 9/137 | 4.50 | 0.00017 | 0.0045 |
| Diabetic cardiomyopathy | 11/137 | 3.73 | 0.00017 | 0.0045 |
| Amyotrophic lateral sclerosis | 15/137 | 2.81 | 0.00028 | 0.0065 |
| Alzheimer disease | 15/137 | 2.67 | 0.00049 | 0.010 |
| Protein processing in endoplasmic reticulum | 9/137 | 3.64 | 0.00082 | 0.016 |
| Longevity regulating pathway - multiple species | 5/137 | 5.60 | 0.0019 | 0.034 |
| **Oliogodendrocyte Precursor Cells (OPCs)** | | | | |
| Steroid biosynthesis | 5/226 | 10.53 | 8.35E-05 | 0.023 |
| Terpenoid backbone biosynthesis | 5/226 | 9.16 | 0.00017 | 0.024 |

SUPPLEMENTAL TABLE 4 KEGG pathways enriched among DEGs in four VTA neuronal subtypes.

| **KEGG Pathways** | **Gene Ratio** | **Fold Enrichment** | ***P*-value** | ***P*adj** |
|---|---|---|---|---|
| **Dopaminergic Neurons** | | | | |
| Synaptic vesicle cycle | 10/250 | 4.82 | 3.81E-05 | 1.13E-02 |
| Parkinson disease | 19/250 | 2.67 | 9.20E-05 | 1.37E-02 |
| GABAergic synapse | 9/250 | 3.85 | 5.23E-04 | 4.45E-02 |
| Folate biosynthesis | 5/250 | 6.80 | 7.19E-04 | 4.45E-02 |
| Neuroactive ligand signaling | 14/250 | 2.68 | 7.50E-04 | 4.45E-02 |
| **GABAergic Neurons** | | | | |
| Prion disease | 32/405 | 2.71 | 2.51E-07 | 7.84E-05 |
| Parkinson disease | 29/405 | 2.52 | 4.12E-06 | 0.00065 |
| Chemical carcinogenesis - reactive oxygen species | 25/405 | 2.59 | 1.17E-05 | 0.00091 |
| Dopaminergic synapse | 18/405 | 3.18 | 1.25E-05 | 0.00091 |
| Pathways of neurodegeneration - multiple diseases | 41/405 | 2.00 | 1.63E-05 | 0.00091 |
| Diabetic cardiomyopathy | 23/405 | 2.64 | 1.94E-05 | 0.00091 |
| Huntington disease | 30/405 | 2.27 | 2.22E-05 | 0.00091 |
| Alzheimer disease | 35/405 | 2.10 | 2.31E-05 | 0.00091 |
| Thermogenesis | 24/405 | 2.40 | 6.03E-05 | 0.0021 |
| Colorectal cancer | 13/405 | 3.51 | 7.16E-05 | 0.0021 |
| Oxidative phosphorylation | 17/405 | 2.90 | 7.38E-05 | 0.0021 |
| Amyotrophic lateral sclerosis | 32/405 | 2.03 | 0.00011 | 0.0028 |
| Relaxin signaling pathway | 16/405 | 2.89 | 0.00012 | 0.0029 |
| Retrograde endocannabinoid signaling | 17/405 | 2.68 | 0.00019 | 0.0043 |
| Lysine degradation | 10/405 | 3.73 | 0.00029 | 0.0058 |
| Autophagy - animal | 18/405 | 2.50 | 0.00030 | 0.0058 |
| RNA degradation | 11/405 | 3.32 | 0.00043 | 0.0079 |
| Growth hormone synthesis, secretion and action | 14/405 | 2.70 | 0.00065 | 0.011 |
| Prolactin signaling pathway | 10/405 | 3.31 | 0.00078 | 0.012 |
| Endometrial cancer | 9/405 | 3.59 | 0.00079 | 0.012 |
| Estrogen signaling pathway | 15/405 | 2.54 | 0.00082 | 0.012 |
| Endocrine resistance | 12/405 | 2.85 | 0.00097 | 0.013 |
| MAPK signaling pathway | 25/405 | 1.96 | 0.00099 | 0.013 |
| Ubiquitin mediated proteolysis | 15/405 | 2.48 | 0.0010 | 0.013 |
| Non-alcoholic fatty liver disease | 16/405 | 2.40 | 0.0010 | 0.013 |
| Protein processing in endoplasmic reticulum | 16/405 | 2.19 | 0.0027 | 0.031 |
| Platelet activation | 13/405 | 2.43 | 0.0027 | 0.031 |
| Inflammatory mediator regulation of TRP channels | 11/405 | 2.61 | 0.0031 | 0.035 |
| AGE-RAGE signaling pathway in diabetic complications | 11/405 | 2.56 | 0.0037 | 0.039 |
| Cholinergic synapse | 12/405 | 2.43 | 0.0038 | 0.039 |
| IgSF CAM signaling | 23/405 | 1.79 | 0.0049 | 0.049 |
| Chemical carcinogenesis - receptor activation | 18/405 | 1.95 | 0.0051 | 0.049 |

SUPPLEMENTAL TABLE 4 (cont.)

| | | | | |
|---|---|---|---|---|
| Chronic myeloid leukemia | 9/405 | 2.75 | 0.0052 | 0.049 |
| **Glutamatergic Neurons** | | | | |
| Oxidative phosphorylation | 25/395 | 4.37 | 3.53E-10 | 1.09E-07 |
| Parkinson disease | 35/395 | 3.11 | 1.73E-09 | 2.68E-07 |
| Prion disease | 33/395 | 2.86 | 4.25E-08 | 4.37E-06 |
| Amyotrophic lateral sclerosis | 37/395 | 2.40 | 5.80E-07 | 4.48E-05 |
| Vibrio cholerae infection | 12/395 | 5.67 | 8.05E-07 | 4.97E-05 |
| Legionellosis | 12/395 | 5.17 | 2.34E-06 | 0.00012 |
| Diabetic cardiomyopathy | 24/395 | 2.82 | 4.00E-06 | 0.00018 |
| Pathways of neurodegeneration - multiple diseases | 41/395 | 2.05 | 8.90E-06 | 0.00034 |
| Apoptosis | 18/395 | 3.17 | 1.35E-05 | 0.00042 |
| Huntington disease | 30/395 | 2.33 | 1.37E-05 | 0.00042 |
| Epithelial cell signaling in Helicobacter pylori infection | 12/395 | 4.07 | 3.06E-05 | 0.00085 |
| Alzheimer disease | 34/395 | 2.10 | 3.30E-05 | 0.00085 |
| Cardiac muscle contraction | 13/395 | 3.60 | 5.54E-05 | 0.0013 |
| Chemical carcinogenesis - reactive oxygen species | 23/395 | 2.44 | 6.62E-05 | 0.0015 |
| Rheumatoid arthritis | 13/395 | 3.30 | 0.00014 | 0.0029 |
| Non-alcoholic fatty liver disease | 17/395 | 2.61 | 0.00027 | 0.0052 |
| Synaptic vesicle cycle | 11/395 | 3.36 | 0.00039 | 0.0070 |
| Kaposi sarcoma-associated herpesvirus infection | 19/395 | 2.34 | 0.00050 | 0.0085 |
| Measles | 15/395 | 2.60 | 0.00063 | 0.010 |
| Thermogenesis | 21/395 | 2.15 | 0.00075 | 0.012 |
| Shigellosis | 22/395 | 2.10 | 0.00082 | 0.012 |
| Collecting duct acid secretion | 6/395 | 5.17 | 0.00085 | 0.012 |
| Cell cycle | 15/395 | 2.29 | 0.0023 | 0.031 |
| GABAergic synapse | 10/395 | 2.71 | 0.0037 | 0.047 |
| Cocaine addiction | 7/395 | 3.44 | 0.0038 | 0.047 |
| Mitophagy - animal | 11/395 | 2.53 | 0.0041 | 0.049 |
| **Unclassified Neurons** | | | | |
| Amphetamine addiction | 13/262 | 6.85 | 3.77E-08 | 1.07E-05 |
| Parathyroid hormone synthesis, secretion and action | 16/262 | 5.06 | 8.94E-08 | 1.27E-05 |
| Circadian entrainment | 14/262 | 5.24 | 3.67E-07 | 3.47E-05 |
| Dopaminergic synapse | 16/262 | 4.37 | 6.86E-07 | 4.87E-05 |
| Glutamatergic synapse | 14/262 | 4.39 | 3.33E-06 | 0.00019 |
| Estrogen signaling pathway | 15/262 | 3.92 | 6.07E-06 | 0.00024 |
| Cocaine addiction | 9/262 | 6.67 | 6.18E-06 | 0.00024 |
| Morphine addiction | 12/262 | 4.79 | 6.69E-06 | 0.00024 |
| MAPK signaling pathway | 22/262 | 2.66 | 2.63E-05 | 0.00083 |
| Endocytosis | 19/262 | 2.74 | 6.52E-05 | 0.0019 |
| Cholinergic synapse | 12/262 | 3.76 | 7.93E-05 | 0.0020 |
| Oxytocin signaling pathway | 14/262 | 3.28 | 9.14E-05 | 0.0022 |

SUPPLEMENTAL TABLE 4 (cont.)

| | | | | |
|---|---|---|---|---|
| Retrograde endocannabinoid signaling | 13/262 | 3.17 | 0.00023 | 0.0050 |
| Protein processing in endoplasmic reticulum | 14/262 | 2.96 | 0.00028 | 0.0052 |
| Longevity regulating pathway - multiple species | 8/262 | 4.69 | 0.00028 | 0.0052 |
| Aldosterone synthesis and secretion | 10/262 | 3.71 | 0.00035 | 0.0062 |
| Spinocerebellar ataxia | 12/262 | 3.03 | 0.00060 | 0.010 |
| GABAergic synapse | 9/262 | 3.67 | 0.00073 | 0.012 |
| Nicotine addiction | 6/262 | 5.32 | 0.00082 | 0.012 |
| Neuroactive ligand signaling | 14/262 | 2.56 | 0.0012 | 0.017 |
| cAMP signaling pathway | 15/262 | 2.41 | 0.0014 | 0.019 |
| Growth hormone synthesis, secretion and action | 10/262 | 2.98 | 0.0019 | 0.024 |
| GnRH secretion | 7/262 | 3.91 | 0.0020 | 0.024 |
| Alcoholism | 13/262 | 2.47 | 0.0024 | 0.028 |
| Purine metabolism | 10/262 | 2.84 | 0.0028 | 0.031 |
| Serotonergic synapse | 9/262 | 2.84 | 0.0044 | 0.048 |
